# The impact of expanded adolescent vaccination against COVID-19 depends on the epidemic status: a mathematical modelling study

**DOI:** 10.64898/2026.08.03.26359567

**Authors:** Anna Fairweather, Ben Swallow, Robyn M Stuart, Cliff C Kerr, Chris Bonell, Russell M Viner, Jasmina Panovska-Griffiths

## Abstract

**Abstract Background/Objectives:** We evaluated the impact of the COVID-19 adolescent vaccination in England at two different epidemic points: the autumn (August-November) 2021, in the presence of a large Omicron epidemic wave, and the autumn (August-November) 2022, when the subsequent Omicron epidemic was at an endemic stage.

**Methods:** Using the Covasim SARS-CoV-2 model for England, under varying vaccine uptake and onset time, we evaluated the impact of a)vaccinating 18+ only versus additional 12+ vaccination from the autumn 2021; and b)the current immunisation strategy at the time versus additional 12+ vaccination from September 2022, projecting the number of new daily SARS-CoV-2 infections, hospitalisations and deaths.

**Results:** In presence of the BA.1 Omicron wave in late 2021, the expanded adolescent vaccination averted 3,000,000 cases across all-ages, 1,010,000 SARS-CoV-2 infections in the period 2-6 months from vaccine onset in the vaccinated cohort. During the Omicron waves in 2022, additional adolescents vaccination did not significantly reduce the COVID-19 burden in the entire population, nor within the vaccinated cohort.

**Conclusions:** Our findings highlight that adolescent vaccination impact depends on the timing/speed of implementation, other present intervention strategies, and the status of the epidemic at the time and it should not be considered as a stand-alone immunisation strategy.

**Highlights:**

- First study to compare the impact of expanding adolescent COVID-19 vaccination at two different epidemic points: one when a large epidemic is spreading and another at a more endemic stage of the epidemic.
- In autumn 2021, in the presence of the newly emerging Omicron variant, adolescent vaccination was necessary to curb infections in both the adolescent cohort and the overall population.
- Earlier implementation at sufficient coverage was more effective in reducing the number of infections at the peak of the Omicron wave, and getting ahead of the epidemic spread.
- In autumn 2022, when the Omicron epidemic was at an endemic stage, adolescent vaccination was not necessary to curb infections in both the adolescent cohort and the overall population.
- Our work shows that implementation, timing and speed of an adolescent immunisation programmes should be considered in conjunction with the epidemic status, incidence and prevalence of the epidemic at that time.

## Introduction

In order to control the spread of SARS-CoV-2 variants in the UK throughout 2020 and 2021 the UK Government imposed three national lockdowns over March to June 2020, November to December 2020 and between January and March 2021. The deployment of effective vaccination against SARS-CoV-2 started in December 2020, and was the planned approach to prevent a further resurgence of COVID-19 in England throughout 2021 alongside the Reopening Roadmap (see Supplementary Materials A). This represented different stages of reopening of communal spaces and reducing restrictions on social mixing following the third national lockdown.

The initial phase of vaccination against SARS-CoV-2 began in December 2020 and targeted elderly people phased by age and high-priority groups such as healthcare workers. Everyone aged 18 years and over, and a small proportion of vulnerable 16-18 years old, were offered a first dose of the vaccine by early July 2021, with the second dose given 8-9 weeks after the first one. In June 2021, vaccination in the UK followed one of two two-dose vaccine regimes approved in the UK: an mRNA-based vaccine produced by Pfizer, and a viral vectored coronavirus vaccine produced by AstraZeneca, with vaccine uptake in adults around 86.8% for the first dose and 67.7% for the second dose in England as of July 18, 2021 [1].

At this point, vaccination was not routinely offered to school-age children and young adults (12-17 years old), apart from a small proportion of vulnerable people within the cohorts. School-age children and young adults have a large number of close interactions that can facilitate COVID-19 transmission. Hence, with the emergence of highly transmissible Omicron BA.1 and BA.1.1 variants in late 2021 and early 2022, adolescents were offered the vaccine, helping to prevent these variants spread. Those aged 16-17 were offered the COVID-19 vaccine from August 23, 2021 and those aged 12-15 were offered the COVID-19 vaccine from September 20, 2021, both as a two-dose regime with the mRNA-based vaccine produced by Pfizer offered 12 weeks apart. As the Omicron variants continued to spread throughout England over 2021, the elderly were offered a booster vaccination from September 2021 to give them additional protection against COVID-19. The scope of this booster vaccine was then widened to include all adults in the UK being offered a booster vaccine at least 3 months after the second dose of the vaccine. As additional Omicron variants emerged in 2022, those aged over 75 were offered a second booster vaccine in the spring 2022. This was further extended to include those aged over 50 from the autumn 2022.

Vaccination against SARS-CoV-2 and its variants is more challenging than, for example, vaccinations against seasonal influenza. Vaccination against seasonal influenza often starts before the onset of infection and before large numbers of infections are evident, whereas vaccination against SARS-CoV-2 was rolled out in a situation of increasing infections. Furthermore, the influenza season in England is of a relatively short duration of a few months, driven by only one viral variant and characterised by a single seasonal epidemic wave. As such it is much easier to prepare and coordinate careful vaccine roll out for seasonal influenza [2]. This is different to the spread of SARS-CoV-2 and the presence of its consecutive variants’ transmission since 2020.

In the literature there is evidence that SARS-CoV-2 vaccination reduces disease severity [3], while there is less evidence on the effect on onwards transmission from SARS-CoV-2 vaccines [4]. Results from the SIREN study in the UK suggest that one dose of the Pfizer vaccine reduces symptomatic cases by between 50% and 70% in those aged 70 years and over, with the second dose improving this protection to between 80% and 90% [5]. Public Health England reported high vaccine effectiveness (VE) after two doses of vaccination against symptomatic disease caused by the Alpha and Delta variants, as well as high levels of effectiveness in preventing hospitalisations and deaths due to COVID-19 [6]. Furthermore, it has been suggested that Omicron is characterised by both a lower initial VE and a faster waning of protection against infection [7]. In contrast, evidence on transmission blocking from COVID-19 vaccines is scarce.

A study of household transmission between fully vaccinated index cases to close contacts in the Netherlands in early 2021 provides evidence that vaccination does offer some protection against transmission for close contacts of vaccinated individuals [8]. However, Eyre et al. suggested that this transmission blocking ability was less effective against the Delta variant than the Alpha variant [9]. Furthermore, the later Omicron variants have been found to have reduced the vaccine’s transmission blocking capability[10].

Hence, there is evidence to suggest that vaccination reduces the symptomatic disease severity but is less good at blocking transmission, particularly against the later Omicron variants. As such, we model the impact of vaccination to be a reduction in disease severity and an increase in the level of neutralising antibodies that an individual has, rather than a directly transmission blocking vaccination.

Mathematical modelling allows simulation of different vaccination strategies to explore the impact of these on future epidemic trajectories. Modelling was widely used to offer scientific advice to policymakers in the UK during the pandemic. Our previous work has considered the effect of the large scale vaccination in early 2021 [11] showing that it was highly impactful and able to significantly reduce the number of SARS-CoV-2 infections from the Alpha and early Delta variant, as well as hospitalisations and deaths, over the period January-July 2021. Other studies have looked at the impact of vaccination over specified time periods, in the presence of different dominating variants and across different settings [12, 13]. These studies are part of the collection of evidence that vaccination has been effective in reducing severe COVID-19 disease and were crucial in reducing the burden of hospitalisations and deaths from COVID-19 over 2020-2022.

During the COVID-19 pandemic, children and adolescents had lower levels of serious disease and death from SARS-CoV-2 [14]. Therefore, they were not included in the initial part of the vaccine strategy for England from December 2020.

This was encouraging to see. However, a vaccination strategy aimed solely at adults would inevitably concentrate infections, and likely transmission, amongst unvaccinated children and teenagers. From June 11, 2021, the COVID-19 infection survey (https://www.ons.gov.uk) suggested an increase in the number of infections in secondary school children and young adults under 30 years old. It was also notable that young people aged 12-29 years had the highest prevalence of virus in the UK before the December 2020 lockdown [15]. The attendance and interaction of primary and secondary school students within schools acts as a bridge between households, contributing to the transmission of the virus [16]. Unvaccinated teenagers could, therefore, be the means of continuing transmission and, whilst at low risk themselves, provide a path for future infection of adults. Balancing all the evidence available at the time, the UK regulator in June 2021 approved the use of the Pfizer-BioNTech vaccine in children aged 12-15 years old and adolescents became a new vaccine target in England from September 2021. At this time, they had high rates of infection and played a larger role in onwards transmission than younger children [17]. Vaccine safety data for this cohort were also published [18].

This paper presents two retrospective analyses undertaken in late 2021 and late 2022 that evaluate the impact of extending the vaccination against COVID-19 to adolescents (12-17 years old) as part of the large scale immunisation strategy in England from September 2021 and September 2022, under assumptions of the vaccine roll out and efficacy reported at the time. The first analysis contrasts the epidemic trajectory during the first Omicron epidemic wave in late 2021 under the vaccine scenario of vaccinating 18+ only and if additionally 12+ were also vaccinated from autumn 2021. The second analysis contrasts the strength of the Omicron wave in late 2022, in the presence of the booster vaccination for the elderly and people at high-risk administered in late 2021 that was then extended to all adults as well as a second booster vaccine given to those aged over 50 in 2022, versus a scenario of additionally offering the booster vaccination to those aged 12+ from September 2022. Across both analyses and immunisation scenarios, we project the number of new daily SARS-CoV-2 infections as well as hospitalisations and deaths related to SARS-CoV-2 over a six-month period from the vaccine roll out start i.e. the periods August 2021 - February 2022 for the first analysis and August 2022 - February 2023 for the second analysis. Our study is the first study to compare the impact of expanding adolescent COVID-19 vaccination at two different epidemic points.

## Methods

We use Covasim, a published individual-based model [19], to simulate the COVID-19 epidemic in England over the period September 2020 to February 2023. Covasim simulates interaction between individuals within four different contact networks: schools, workplaces, households, and community settings. Our work continues chronologically on the work of Panovska-Griffiths et al. [11, 20], and focuses on evaluating different immunisation strategies for 12-17 years old in late 2021 and late 2022. For these analyses, we start with a baseline model that was calibrated to simulate the epidemic in England (see Figure 1). The details of the calibration and the model parameters can be found in Supplementary Materials B and Supplementary Materials D, respectively. The calibrated model is then amended to investigate different adolescent vaccination strategies.

**Figure 1:**
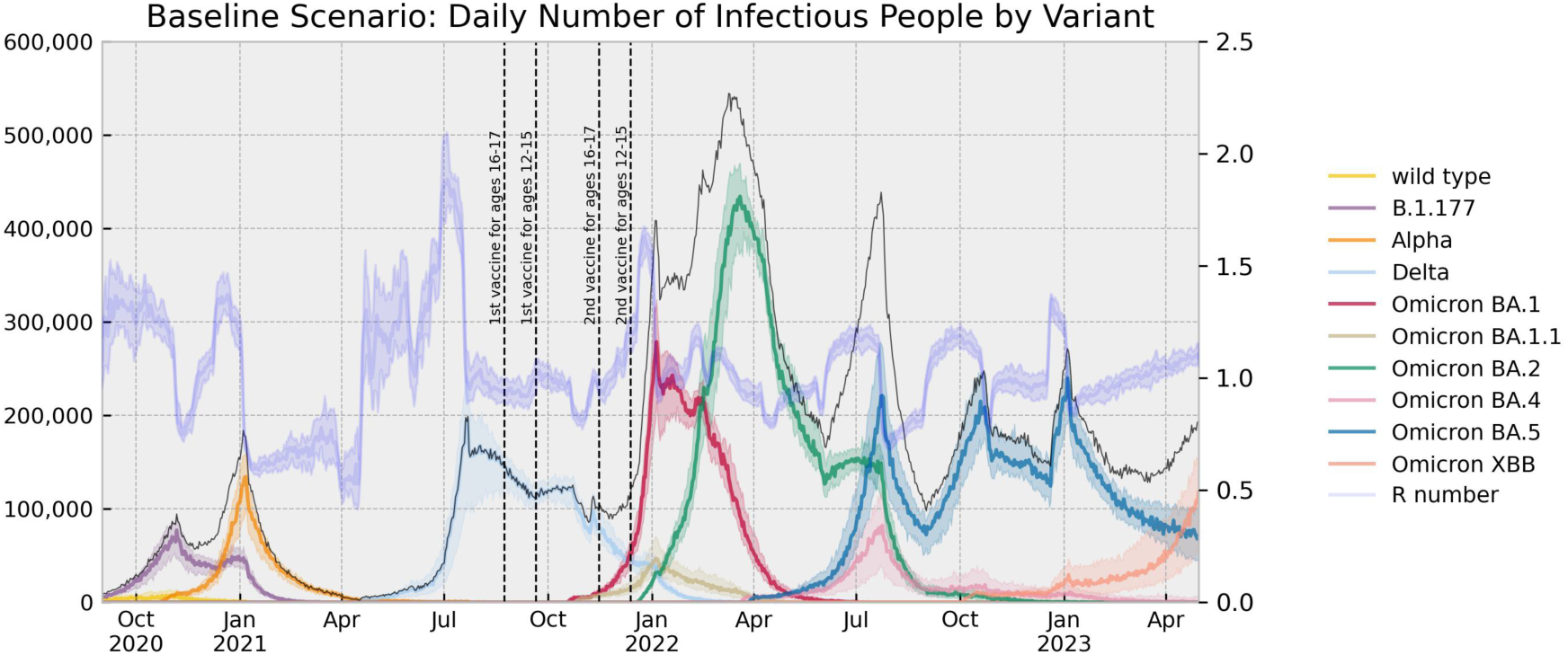
Projection of the daily number of infectious people by variant and the effective reproduction number produced by the baseline scenario to simulate the spread of the COVID-19 epidemic in England between September 2020 and April 2023.

### Modelling different immunisation strategies

Vaccine efficacy is modelled through an immune response that primes and boosts neutralizing antibodies (NAbs) in individuals and then relates the level of NAbs to protection against infection, symptomatic disease and severe disease. The model accounts for waning of NAbs over time and has been fitted to vaccine efficacy and effectiveness trials data [21]. For this study, we model the reported immune response induced by the Pfizer/BioNTech and Oxford/AstraZeneca vaccines.

### Data sources

We calibrate the model to publicly available data from the UK COVID-19 dashboard over the period January 2020 to January 2023 (https://coronavirus.data.gov.uk). We also use data from the publicly available COVID-19 Genomic Surveillance Data from the Sanger Institute (https://covid19.sanger.ac.uk/lineages/raw) to include the estimated proportion of infections belonging to a variant during each week (see Supplementary Materials C.1 for visualisation). We use this data to see when variants emerged and how they interact with one another.

Our final source of data is the Coronavirus Infection Survey: England, from the Office of National Statistics (https://www.ons.gov.uk). This gives an estimate for the number of newly infectious people over the period of a week. We adjust these to give the number of infectious people per day (see Supplementary Materials D.1 for details).

### Calibration

By using the baseline calibrated model, and version 3.1.4 of Covasim, we run 30 simulations and generate the median of these simulations as the central estimate as well as the 25th and 75th percentiles. Across the different scenarios, we project the number of hospitalisations, COVID-19 diagnoses, deaths and infections due to COVID-19 over the study period, as in Figure 2 for the baseline model. Using the calibrated model, two separate analyses are undertaken (parametrisation details for these are found in Supplementary Materials D.2).

**Figure 2:**
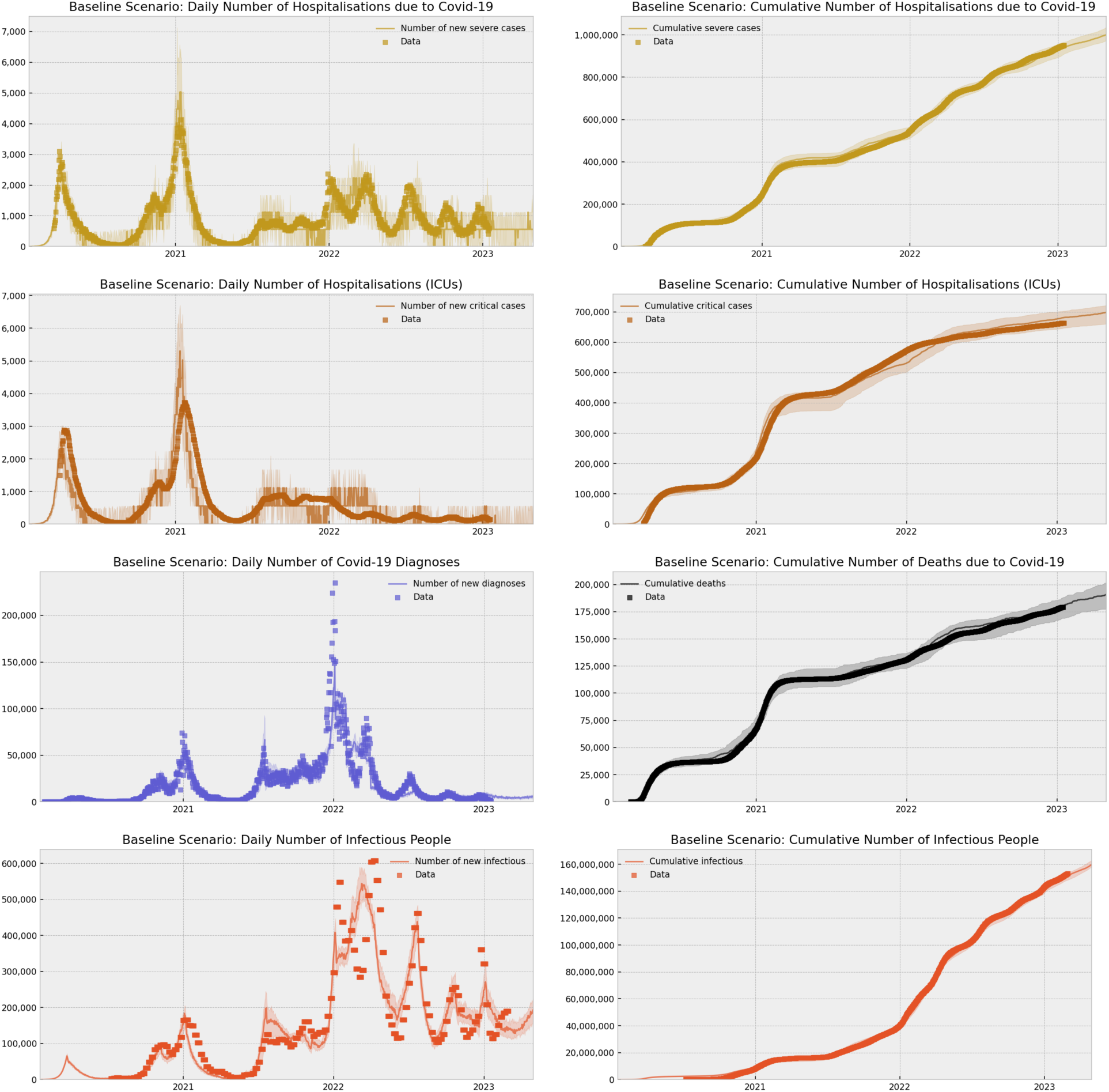
Comparison of the results of our baseline scenario simulated in COVID-19 and the observed data. The data is sourced from https://coronavirus.data.gov.uk and https://www.ons.gov.uk. Our projections simulate: a) the daily number of hospitalisations due to Covid-19 in England, b) the cumulative number of hospitalisations due to Covid-19 in England; c) the daily number of ICU admissions due to Covid-19 in England; d) the cumulative number of ICU admissions due to Covid-19 in England; e) the daily number of reported positive diagnoses of Covid-19 in England; f) the cumulative number of deaths due to Covid-19 in England; g) estimated and simulated daily number of infectious people in England; and h) estimated and simulated cumulative number of infectious people in England.

### Modelling the impact of varying adolescent vaccination in autumn 2021

The first analysis explores the impact of adolescent vaccination on the Omicron epidemic in England over August 2021 - February 2022. A large-scale study of the vaccine uptake across the UK nations [22] showed that, in England, 34% of 5-17 year-olds received their first vaccine, 20% received their second vaccine, and 2% received their booster, leaving 66% of 5-17 years old unvaccinated between 4th August 2021 and 31st May 2022. In the UK, among the age group we have focussed our modelling on the highest uptake was in the 16–17 year-olds: 71% received their first vaccine, 54% received their second, and 14% received their booster. Lower uptake was observed in 12–15 year-olds: 59%, 39% and 1%, respectively.

Aiming to cover as many scenarios as possible, and using these reported statistics, we simulated four vaccine uptake scenarios: 0%, 50%, 70% and 90% of adolescents vaccinated. These scenarios reflect instances of not rolling out the vaccination in adolescents (0%), an uptake similar to the average two-dose vaccine uptake in adolescents (50%), uptake reflective of the highest uptake which was in 16-17 year-olds (70%), and an extremely high vaccine uptake similar to that of the current at the time MMR vaccine (90%). We note that at the time of advising stakeholders on the impact of adolescent vaccination in July 2021 we only modelled the scenarios of 0% and 50% vaccine uptake. We are now including two additional vaccine uptake scenario based on the statistics in [22].

For simplicity, when simulating our four adolescent vaccine uptake scenarios, we implemented the same % uptake in those aged 12-15 and 16-17 even though clear differences were observed in the data. For each of these adolescent vaccine coverage scenarios, we delayed the start date of the vaccine roll out by two four-week intervals, starting vaccination in early August, September and October. A summary of these 12 scenarios can be found in Table 1.

**Table 1:** Scenarios used to model the impact of varying adolescent vaccination in autumn 2021. All scenarios involve the same projection as the baseline scenario up to the point of adolescent vaccination, then they simulate varying adolescent uptake starting at different time points for the period August 2021 - March 2022.

|  | 0% | 50% | 70% | 90% |
| --- | --- | --- | --- | --- |
| August 2021 | 0% adolescents vaccinated | 50% adolescents vaccinated, starting vaccine roll out in August 2021 | 70% adolescents vaccinated, starting vaccine roll out in August 2021 | 90% adolescents vaccinated, starting vaccine roll out in August 2021 |
| September 2021 | 0% adolescents vaccinated | 50% adolescents vaccinated, starting vaccine roll out in September 2021 | 70% adolescents vaccinated, starting vaccine roll out in September 2021 | 90% adolescents vaccinated, starting vaccine roll out in September 2021 |
| October 2021 | 0% adolescents vaccinated | 50% adolescents vaccinated, starting vaccine roll out in October 2021 | 70% adolescents vaccinated, starting vaccine roll out in October 2021 | 90% adolescents vaccinated, starting vaccine roll out in October 2021 |

### Modelling the impact of different booster vaccinations in autumn 2022

Our second analysis explored the impact of an additional adolescent booster vaccine during the Omicron epidemic in England over August 2022 - February 2023. A summary of these scenarios can be found in Table 2. We first use the booster vaccine strategy that is already in our baseline scenario which replicates the strategy followed in England. Following this, we run another scenario with the same strategy followed by an additional adolescent booster vaccine which includes a single dose of a booster vaccine for those aged 12-18 starting from September 2022. We simulate a similar proportion of adolescents receiving the booster as the proportion of adolescents who received the two-dose vaccination in the baseline scenario.

**Table 2:** Scenarios used to compare the impact of an adolescent booster vaccination starting in the Autumn of 2022 (see Supplementary Materials D.3 for further details. Both scenarios share the same parameters with the exception of the additional booster vaccine being included in the Adolescent Booster Scenario, then these scenarios are projected for the period August 2022 - March 2023.

| No Adolescent Booster Scenario | Adolescent Booster Scenario |
| --- | --- |
| Baseline scenario | Baseline scenario with an additional booster vaccine for adolescents starting 01/09/2022 |

## Results

### The impact of expanding the adolescent vaccination in England on the epidemic over August 2021- February 2022

Following our calibration of the Omicron variants, we looked at varying the adolescent vaccination roll out whilst keeping the rest of the model parameters the same. As expected, and as Figure 3 demonstrates, the epidemic is increasingly more controlled at the end of 2021/beginning of 2022 as the uptake of vaccines in adolescents increases. Independent of when we start adolescent vaccination, we can see a clear reduction in the daily number of infectious people when we compare the 0% adolescents vaccinated scenario to the other scenarios that include adolescents’ vaccination. For example, our simulation projects an additional (median) 20,100 newly infectious people per day by January 1, 2022 when we start adolescent vaccination from August 2021 and compare the 0% and 50% vaccine uptake scenarios. We see a further a reduction in the median number of newly infectious people of 67,900 when we look at the 70% uptake scenario compared with 50% uptake scenario. However, increasing the uptake further to 90% does not have the same effect, with only a small additional reduction in the median number of newly infectious people of 2,500. This may be a result of the fact that only marginally more people are incrementally vaccinated with this additional maximum uptake. Overall, and across all uptake levels, over the period August 2021-February 2022, our findings show that the vaccination of adolescents clearly reduces the number of infections during the early emergence of Omicron variants in late 2021.

**Figure 3:**
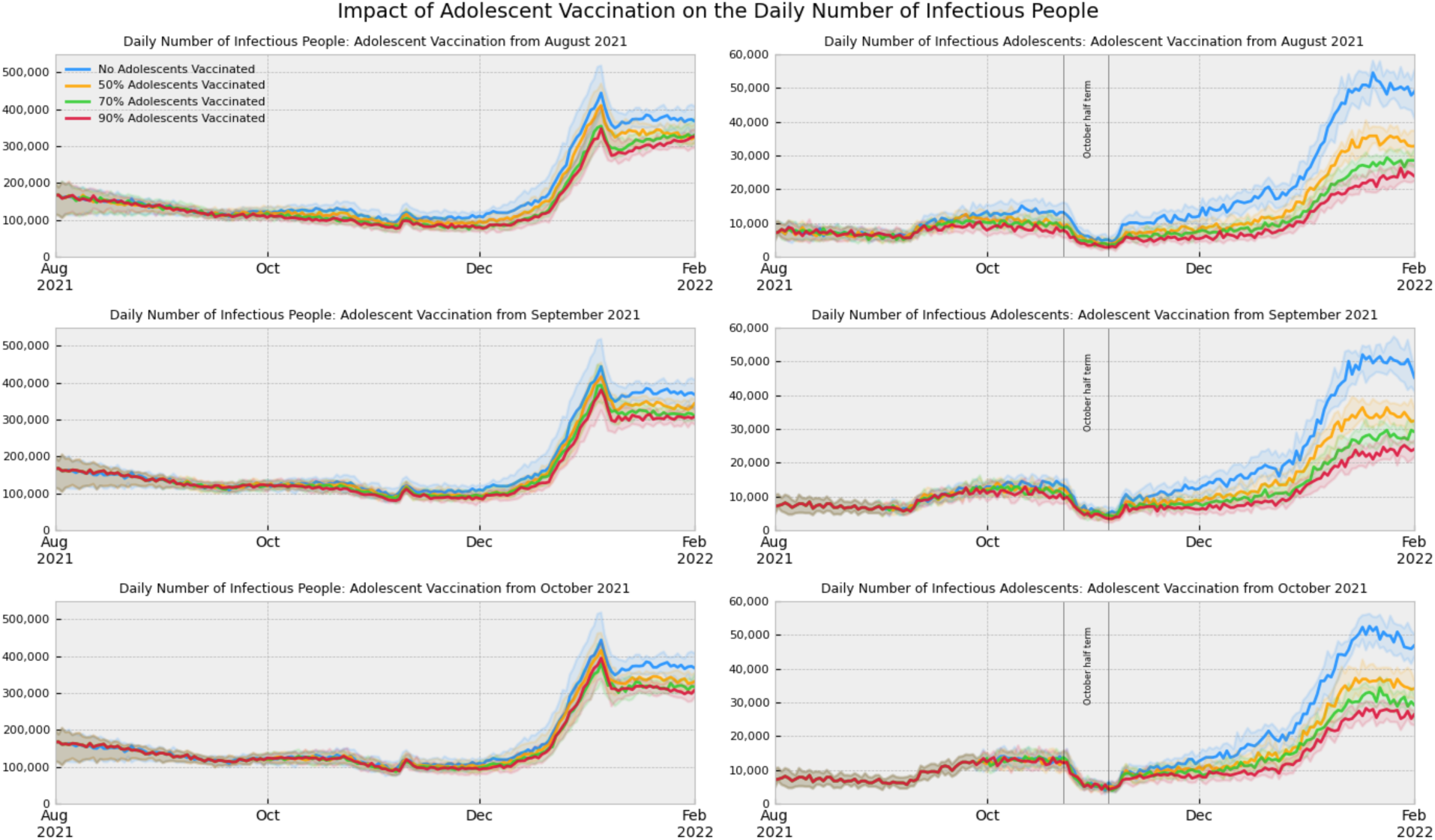
Comparative graphs showing the impact of varying adolescent vaccine uptake on the spread of the COVID-19 epidemic in England through the number of infectious people in the whole population and in the target population (adolescents), and how this changes when vaccination is started in early August, September or October 2021.

Interestingly, there is a more notable reduction in the number of cases in the scenarios when adolescent vaccination is rolled out earlier. When adolescent vaccination is rolled out in early August, there is a clear decrease in the cumulative number of infections across the whole population as the percentage of vaccine coverage in adolescents is increased. When this vaccine roll out is delayed by four/eight weeks, we see less distinction in the number of infections across the whole population when adolescent vaccination is increased to 70% and 90%. However, when we look at the adolescent population alone we see similar reductions in infections as vaccination uptake increases regardless of vaccine rollout start date. This indicates that introducing the vaccine roll out earlier would help to reduce the strength of the Omicron wave amongst the whole population, as the peak immunity amongst adolescents would coincide with the peak of the Omicron wave in early 2022. This early vaccination would have limited impact on the number of infections amongst adolescents themselves, instead protecting the wider population through a higher population level immunity at the peak of the wave. Hence, our results indicate that if we can vaccinate early enough then having a higher percentage of adolescents vaccinated would be beneficial. If we start vaccination later in October 2021, however, there is no improvement gained from increasing adolescent vaccine uptake to 90% from 70% as there is no reduction in the daily number of newly infectious people at the peak of the Omicron wave. In comparison, if we start vaccination in August 2021, there is some benefit in reducing the number of daily infections, although this remains small.

Overall, these results show that the interplay between timing of the vaccination and the uptake level is important. For example, at the peak of the Omicron wave there were 39,100 fewer daily infections in the 70% uptake scenario starting in August 2021 when compared to the 90% uptake scenario starting in October 2021. In contrast, however, when we compare the 50% uptake scenario starting in August 2021 with the 70% uptake scenario starting in October 2021 we find that vaccinating early did not reduce the number of daily infections, and instead we saw 28,800 more daily infections in the 50% uptake scenario starting in August. These results suggest that we need sufficient coverage regardless if we start vaccination in August or October. We find that there is benefit in increasing vaccine uptake to 70% even if we do not start vaccine rollout early. However, starting early is still better, as shown by comparing the 70% uptake with the August start with the 90% uptake with the October start. This reveals that having an earlier vaccine rollout start with less vaccine coverage is more impactful than starting vaccination later but with higher vaccine coverage, as long as the coverage in both scenarios is sufficient. This comparison shows that we need to start vaccination early before the epidemic takes over to prevent the most infections.

Figure 4 further shows the impact of this vaccine delay. While there is a reduction in the cumulative number of infections across all scenarios, a more notable reduction is present in the early vaccine roll out scenarios at the peak of the Omicron wave. When comparing the 0% and 50% uptake scenarios on January 1, 2022, our simulation projects a reduction in the median cumulative number of infections of 1,980,000 when we start vaccination from August 2021. We see a further reduction in the cumulative number of infections of 880,000 when we compare the 70% scenario to the 50% scenario, and 270,000 when we compare the 90% scenario to the 70% scenario. In contrast, when comparing the 0% and 50% scenarios we only see a median reduction of 1,530,000 if we start vaccination from September 2021 and 617,000 if we start vaccination from October 2021. These results confirm the importance of starting adolescent vaccination early, ahead of the epidemic wave, and with sufficient coverage. We did not observe similar patterns of reduction in the number of hospitalisations or deaths, likely due to the fact that these disease outcomes only marginally effect adolescents directly. Additional figures relevant to these projections can be observed in Supplementary Materials C.2.

**Figure 4:**
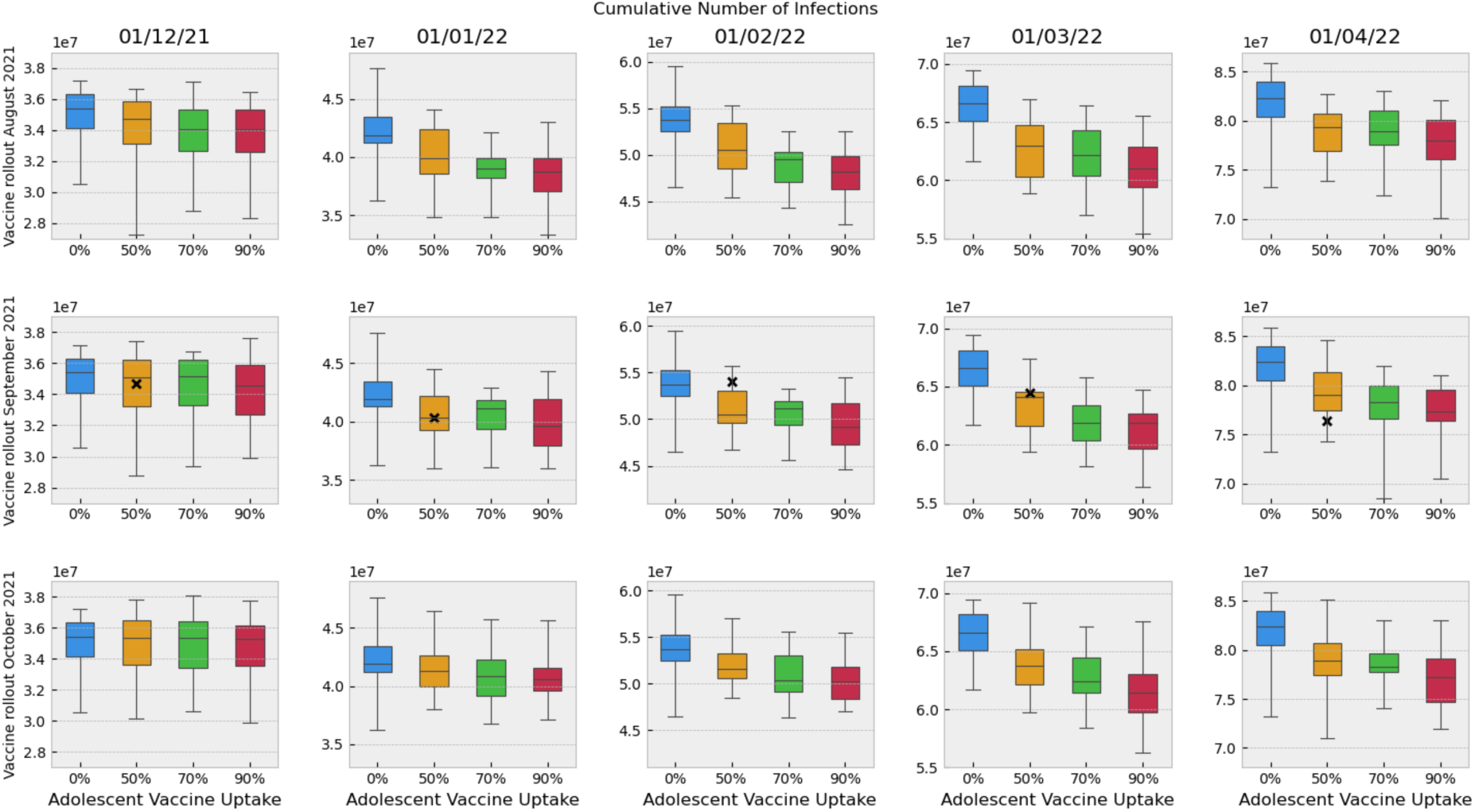
Box plot showing the impact of varying adolescent vaccine uptake on the cumulative number of COVID-19 infections in England and how this changes when vaccination is started in early August, September or October 2021. The x indicates the estimated cumulative number of infections in England and projected onto the adolescent vaccine strategy modelled which most closely replicates the actual vaccine roll out in England.

We have also created a 3D surface to show how the cumulative number of infections varies with % of adolescents vaccinated and vaccine roll out start date (see Supplementary Materials C.3). We have taken the 3D slice on March 1, 2022. This date has been chosen to account for high VE around 4-6 months after the first vaccine is administered. As expected, it is observed that increasing adolescent vaccine uptake reduces the cumulative number of infections in the whole population. Furthermore, reducing the delay in vaccination, in general, also reduces the total number of infections. This again suggests that if we had vaccinated early enough so that the peak level of COVID-19 immune resistance amongst adolescents would be at the same time as the Omicron wave peak then we would have achieved the most significant reduction in Omicron peak size.

To better understand the impact of the adolescent vaccine roll out, we can look at the simulated projection within the target population only (ie. adolescents). As Figure 3 shows, there is a significantly larger proportional reduction in the number of infections averted among the targeted adolescents cohort than across the whole population. Adolescents account for 6,430 of the 20,100 (median) daily additional infectious people by January 1, 2022 when we compare the 0% and 50% vaccine uptake scenarios and start adolescent vaccination from August 2021. This is a much larger proportion (31.9%) than the proportion of adolescents in the whole population (*≥* 7%). This suggests that the biggest impact of the expanded vaccination to adolescents in the autumn 2021 was the targeted adolescents’ group. Our results here align with our general expectation around vaccine delivery: the cohort who receive the largest direct benefit from vaccination is the vaccinated cohort itself. This is especially true given that the vaccine is not modelled as having specific transmission blocking properties, but rather increases the number of neutralising antibodies that an individual has.

### The impact of expanding the adolescent vaccination in England on the epidemic over August 2022-February 2023

We find that simulating a booster vaccine for adolescents in the autumn 2022 in England projects a much smaller reduction in the number of infections across the whole population (see Figure 5) when compared with the impact of the two-dose adolescent vaccination. We considered the period from 2 months to 6 months after adolescent booster vaccine roll out began (November 1, 2022 - March 1, 2023) and found the (median) reduction in the cumulative number of infections over this period with an adolescent booster vaccine present to be 1,220,000. In comparison, the same period in the previous year (November 1, 2021 - March 1, 2022) saw a (median) reduction of 2,340,000 infections (a 91.5% increase on the cumulative number of infections avoided) in the presence of realistic at the time (50%) adolescent vaccine coverage compared to no adolescent vaccination.

**Figure 5:**
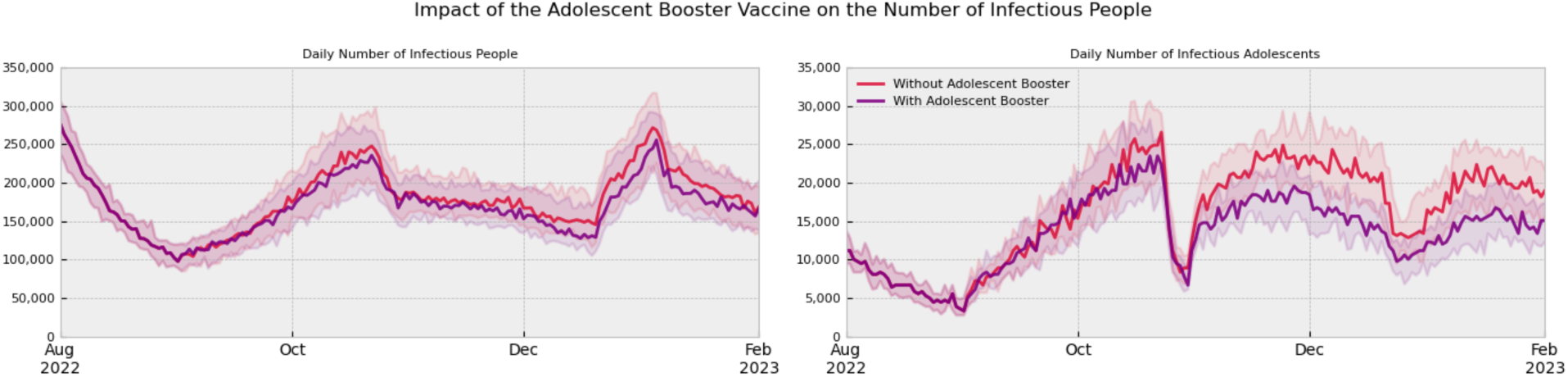
Comparative graphs showing the impact of a single dose of the booster vaccine for adolescents starting in September 2022 on the number of infectious people in the whole population and in the target population (adolescents).

This adolescent booster vaccine is simulated in the presence of the elderly and high-risk booster vaccine roll out in spring 2022. We simulate a similar proportion of adolescents receiving the booster as the proportion of adolescents who received the two-dose vaccination in the baseline scenario. As described and shown in Figure 5, the reduction in the number of infectious adolescents is also small and not significant. In the respective same four month period (November 1, 2022 - March 1, 2023) we see a (median) reduction in the cumulative number of infections in adolescents of 536,000 when we include the adolescent booster vaccine. Considering this against the same period the previous year (November 1, 2021 - March 1, 2022), we saw a (median) reduction of 1,010,000 adolescent infections when we compared the 0% vaccine uptake with the baseline scenario. The booster vaccine scenario has almost half as large a reduction in the cumulative number of adolescent infections during the same four month period a year later, and a similar pattern is reflected in the whole population. This is visualised further in Figure 6. Additionally, there is no notable impact on the number of hospitalisations or deaths in the whole population and the adolescent proportion of the population (see Supplementary Materials C.4).

**Figure 6:**
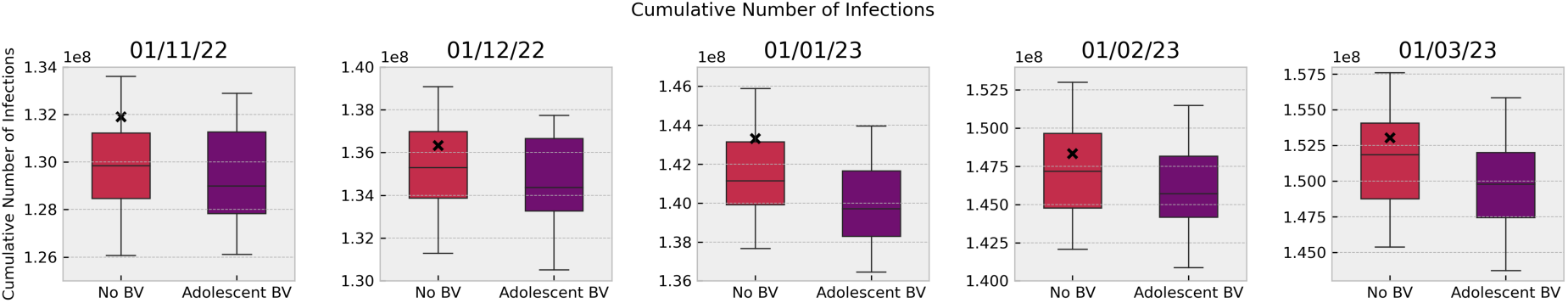
Box plot showing the impact of a single dose of the booster vaccine for adolescents starting in September 2022 on the cumulative number of infections in England. The x indicates the estimated cumulative number of infections in England and is compared to the no adolescent booster vaccine strategy modelled which most closely replicates the actual vaccine roll out in England.

## Discussion

We have presented the results of our retrospective modelling analyses undertaken over autumn 2021 to winter 2022 and autumn 2022 to winter 2023 that provided scientific evidence to policy decision makers for the impact of the adolescents’ immunisation strategies in England. We use the stochastic individual-based model Covasim, calibrated for the English epidemic between January 2020 to April 2023, to model the transmission of different SARS-CoV-2 variants over August 2021-April 2023 and simulate different scenarios. We contrast vaccinating 18+ only versus additional 12+ vaccination in the autumn 2021 and the autumn 2022 in England.

Our results show that in the presence of the highly transmissible and emerging BA.1 Omicron wave in late 2021, expanding the vaccination to 12-17 years old in the autumn 2021 led to significant reduction in cases: from a median of 54,000,000 cumulative infections on 01/02/2022 in the no-adolescent vaccination scenario, to 51,100,000 infections in the Baseline Scenario (which replicates the actual roll out of adolescent vaccination in England). A large proportion of the benefits were observed within the vaccinated 12-17-year-old cohort; over 1 million additional infections were averted in adolescents in the period from 2 months to 6 months post adolescent vaccine roll out (November 1, 2021 - March 1, 2022) when adolescent vaccination was simulated.

In contrast, during the later Omicron waves in the autumn 2022 in England, and in the presence of the booster vaccination of elderly/high-risk people, additional vaccination of 12-17 years old from August 2022 did not lead to such a large reduction in the number of infections amongst adolescents: less than 550,000 additional infections were averted in adolescents in the period from 2 months to 6 months post adolescent booster vaccine roll out (November 1, 2022 - March 1, 2023) when we included an adolescent booster vaccine. Furthermore, over this period, additional adolescents vaccination did not lead to significant reduction in COVID-19 hospitalisations or deaths in the entire population or within the vaccinated adolescents cohort.

### Novelty of findings

#### Our study has many novelties

Firstly, these analyses are the first study to compare the impact of expanding adolescent COVID-19 vaccination at two different epidemic points: one when a large epidemic is spreading and another at a more endemic stage of the epidemic. For the first time, we show that expanding the COVID-19 vaccination to include adolescents can have a different impact at different epidemic points. In the autumn 2021, in the presence of the highly transmissible and emerging Omicron variant, adolescent vaccination was necessary to curb infections in both the vaccinated cohort and the overall population. A year later, in the autumn of 2022 when the subsequent Omicron epidemic was at an endemic stage, and the spreading Omicron variant was less transmitted due to a higher level of background population immunity, adolescent vaccination was not necessary. Hence, our work illustrates that vaccination of adolescents against COVID-19 should be considered in conjunction with the epidemic status, incidence and prevalence at the time, as well as other vaccination strategies rolled out at the time, and also possible side effects.

Myocarditis has been identified as a rare but significant adverse event associated with COVID-19 vaccination. Those at the highest risk of myocarditis are men aged 12-24 who receive a second dose of the COVID-19 mRNA vaccine [23]. Given this age category includes adolescent males, this adverse reaction, along with other potential side effects, should be considered in conjunction with modelling evidence, such as this study, on the impact of adolescent vaccination when policy recommendation are being made to ensure that an all-composing approach in deciding vaccination strategy is made.

Secondly, our results show that adolescent vaccination in the autumn 2021, as the first Omicron wave was emerging, had a large impact on the number of COVID-19 infections. A significant proportion of the reduction in the number of COVID-19 infections was observed in the vaccinated target cohort, with less of an impact on hospitalisations and deaths related to COVID-19. This agrees with existing published studies [24, 25, 26, 27]. However, novel to these studies we were also able to contrast this to an analogous period the following year, showing that during endemic COVID-19 period, in contrast, adolescent vaccination has less impact. This has not been shown in previous work. .

Thirdly, our study is the only one to date to consider long-period of SARS-CoV-2 transmission, and specifically compare two consecutive years’ dynamics. Existing studies have looked at the impact of vaccination of adolescents and children in England [26, 27], the United States of America [24] and the Netherlands [25] at only one time point in the epidemic. We have instead focused on one setting (England), but consider two analogous time periods over two consecutive years, hence allowing us to explore longer time dynamics and hence give a more informative policy advice for future pandemic preparedness.

Fourthly, our study is the only study to date that can capture consecutive SARS-CoV-2 variants and incorporate their cross-variants dynamics. None of the existing models incorporated different variants and, hence, could not model their cross-immunity. This is something that, to date, only the Covasim model has been able to incorporate. In [27] there was a discussion of a new variant coming, which was the first Omicron wave, in November 2021, but this was not modelled explicitly. Capturing multiple variants, over long time dynamics allowed us to give a full picture of the COVID-19 epidemic over the entre pandemic period in a setting, something that has not been done.

Fifthly, as well as discussing, for the first time, the implementation of an adolescent vaccination over a whole course of an epidemic wave, we were able to generate results on the interplay between timing and speed of this implementation. Our results suggest that vaccinating adolescents early at a sufficient uptake level is key to reducing the number of infections at the peak of the Omicron wave, and getting ahead of the epidemic before it takes over. This is important pandemic preparedness advice that is useful for both pandemic strategies planning as well as capacity planning of the rollout of future potental adolescent vaccination programmes.

And finally, our study is the only one to date that can dynamically capture vaccine or infection waning. This allowed us, here, for the first time, to explore the effects from such waning on longer term SARS-CoV-2 dynamics when considering vaccination over a longer time period. This is especially relevant as we consider longer term simulations e.g. 4-6 months after the vaccine start. The interplay between vaccine/infection protection and waning is an important aspect to include, and our model and this analysis is the first to be able to do this.

#### Strengths of the modelling work

As well as these novelties, our study with the Covasim model has a number of additional strengths. Firstly as a stochastic model, Covasim is able to capture uncertainty in the predicted trajectories emerging from both the inherent stochasticity and the uncertainty around the parameters values. Our results were based on taking the median of 30 simulations from a stochastic process, but we note that the results and conclusions drawn did not change when we ran more simulations. We also note that this uncertainty increases when predictions are made over a longer time period. While it is important for us to calibrate the model for the long time frame (2020-2023) to see the impact of vaccine scenarios, we are mindful that projecting results of any model, including ours, too far into the future based on current data is unwise due to the uncertainty about the continued validity of model assumptions.

Secondly, Covasim has the ability to readily capture different ways of modelling vaccination as well as different cohorts being targeted for vaccination. This allowed us to readily design scenarios that contrast the impact from different vaccine strategies on both adolescents as well as other population cohorts. When we examine the projected impact in the adolescents versus the whole population, unsurprisingly, we find that that the individuals who benefitted most from extending vaccination were adolescents themselves, with the benefit highest in the presence of a large epidemic wave. This is due to the indirect protection against infection and the direct protection against severe disease provided by vaccination in that cohort, which is emphasised during a large epidemic wave. In general agreement with published literature, we modelled vaccination as an increase in the level of neutralising antibodies in an individual rather than modelling the vaccine as transmission blocking. Hence, the vaccine reduces an individuals chances of becoming symptomatic and severely ill. Indirectly, this reduces the probability that an individual transmits the virus and is in agreement with literature.

And thirdly, with Covasim we can readily capture the spillover effect from vaccinating adolescents on the wider population e.g. contrast the reduction in disease outcomes in adolescents vs the general population. While we do observe some reductions in disease outcomes in the whole population, particularly over the large epidemic wave in 2021, this is much less. Again, this is expected as we do not model an effect from vaccination in preventing onward transmission directly, but through a reduction in the probability that an individual is symptomatic or gets severe disease (due to COVID-19). The impact on COVID-19 hospitalisations was also less in all scenarios studied. As a result, we conclude that if the aim of policy, as it was in England throughout 2020 and 2021, is to protect the elderly and not overwhelm the NHS i.e. the hospitals in England, then vaccination of adolescents and children may not be the most effective policy decision. Previous studies have highlighted that, since adolescents have large number of contacts [28], reduced social mixing and social distancing measures can be an effective strategy to reduce COVID-19 infections, specifically if they target the age groups that contribute most to secondary spread [29].

### Limitations of the work

We note as this is a modelling study, it has a number of limitations. Firstly, the analyses presented here are retrospective and were initially performed in the summer 2021 and summer 2022. Hence the model assumptions mostly reflect the available knowledge at that time, although we have updated some of this knowledge in the calibration process; specifically the information on the characteristics of the later Delta and the Omicron variants, which were not known at the initial run of the analysis. Secondly, we used assumptions on both the vaccine efficacy against onward transmission across Delta and Omicron variants, and the waning protection from both vaccination and infection. Specifically, we assumed a single antibody waning function for all individuals and all types of immunity, with individual- and immune-level variation in the level of NAbs. This can be explored further in future work, where we consider the difference between short and long term dynamics. For example, differences in the antibody kinetics of natural versus vaccine-derived neutralising antibodies are relevant in short-term dynamics, while long-term dynamics are driven primarily by the introduction of new immune-evading variants. Exploring aspects like these, with layers of different vaccination strategies is part of our planned future work but is out of scope for this paper. Thirdly, we calibrated the model to aggregated national data on reported cases, hospitalisations and deaths from England, whilst checking that the fits are good in the adolescent cohort we are studying. Fitting more granular distribution of these epidemic metrics across different age groups is possible, but since we are considering the impact of the adolescent immunisation on the whole population, this is sufficient for this study. Fourthly, we calibrated our model using the Optuna hyperparametrisation to search a set of optimal parameters that gave a good fit of the model projections to the available data over the long period of study. We note that as with any calibration, our baseline model tries to reflect the data as accurately as possible but is not able to perfectly match every data point. For example, we have a very good fit of the model projections to the genomic data over the period of the emergency of the Alpha and early Omicron variants, while the fit is worse over the period of the emergency of the Delta or the later Omicron variants. Specifically, we note the discrepancy between the model infection estimates and the officiant variants data over the period of re-emergence of the BA.2 Omicron variant alongside the XBB variant in early 2023. This has resulted in a worse fit to the variant distribution data from the Sanger Institute. Despite this, overall the prominence of the variants in circulation within the baseline model is similar enough to that estimated by the Sanger Institute genomic data, and hence we do not expect any discrepancies to impact our results.Lastly, while we included non-pharmaceutical interventions (NPIs), such as test-trace-isolate (TTI) strategies, these were based on data from England and we did not explicitly explore the interplay between NPIs and vaccines. In reality, if the number of severe disease outcomes increased rapidly and overwhelmed the healthcare systems, NPIs could be rapidly enforced. In light of this, we note that the projections we show here are one possible outcome of disease outcome projections.

### Conclusions

In summary, our application of a stochastic agent-based model shows that the expansion of the immunisation strategy in England to include adolescents in the autumn 2021 was necessary to curb infections across the whole population and in the target cohort as the first Omicron wave was emerging. Furthermore, earlier vaccine implementation led to a higher reduction in COVID-19 burden. However in the autumn of 2022, in the presence of later Omicron waves and when the epidemic was at an endemic stage, the roll out of booster vaccines for all adults and a second booster vaccine for the elderly and high-risk people was sufficient to curb infections, hospitalisations and deaths from COVID-19. Expanding the booster vaccination programme to include adolescents at this stage did not seem to have additional benefits. At both time points, the impact from vaccinating adolescents was mostly in preventing infections in the target group, with less pronounced impact on infections in the whole population, or hospitalisations and deaths related to COVID-19 across the population or the target group.

By showing that the impact of adolescent vaccination against COVID-19 depends on the epidemic status, how quickly the roll out can be implemented, and what other strategies are present at the time, our findings suggest that adolescent vaccination should not be considered and evaluated alone, but in conjunction with other strategies and the epidemic status at the time.

## Declarations

### Ethics approval and consent to participate

All methods were performed in accordance with the relevant guidelines and regulations for human research as listed in the Declarations of Helsinki. The study doesn’t require ethics approval.

### Availability of data and material

The numerical code and the data used to generate the projections across the scenarios reported in this paper are available at https://github.com/spet5215/Adolescent_vaccination_analysis.

### Authors Contributions

JPG and RMV conceived the study with support from CB. JPG and AF developed the specific Covasim modelling framework for this study based on responsive modelling work by JPG in 2021 and 2022. JPG, RMV and CB defined the different scenarios in the UK context following conversations with the UK Health Security Agency (UKHSA), Scientific Pandemic Influenza Modelling Operational Group (SPI-M-O) and Scientific Pandemic Influenza Behaviour Group (SPI-B) and Scientific Advisory Group for Epidemics (SAGE) which give expert advice to the UK Department of Health and Social Care and wider UK Government. AF ran the modelling scenarios. AF and JPG wrote the manuscript with input from BS, RMS, CCK, CB and RMV. All authors approved the final version. JPG is the manuscript’s guarantor.

## Data Availability

All data produced in the present study are available upon reasonable request to the authors.

https://covid19.sanger.ac.uk/lineages/raw

https://www.ons.gov.uk

https://coronavirus.data.gov.uk

## Acknowledgements

JPG’s work is supported by the UK Health Security Agency and the UK Department of Health and Social Care. AF is supported by a PhD studentship from the UK Health Security Agency as part of the Oxford EPSRC Centre for Doctoral Training in Healthcare Data Science (EP/Y035321/1). The funders had no role in the study design, data analysis, data interpretation, or writing of the report. The views expressed in this article are those of the authors and not necessarily those of the UK Health Security Agency, the UK Department of Health and Social Care or the EPSRC Centre for Doctoral Training in Healthcare Data Science.

