## Supplementary Material for "The impact of expanded adolescent vaccination against COVID-19 depends on the epidemic status: a mathematical modelling study"

#### A Reopening roadmap

The reopening roadmap began with Stage 1: the reopening of schools on March 8, 2021 and the relaxation of the 'stay at home rule' on March 29, 2021. Stages 2 and 3 followed from April 12, 2021 and May 17, 2021 – with further aspects of society relaxed slowly. Stage 4 of the Reopening Roadmap, which was originally planned from June 21, 2021, was delayed to July 19, 2021 as a consequence of the increase in the number of COVID-19 cases from the middle of April 2021. This was attributed to the emergence and spread of the more transmissible Delta variant from late March 2021.

#### B Baseline model

Here we give a concise overview of the structure of Covasim and how we developed our Baseline model. We refer to [19] for a full explanation of the Covasim model.

A single run of Covasim follows a logical sequence of events that begins by creating a simulation object with a population of agents and then assigning each agent an age, sex and health state. The health state of an agent relates to predefined states of COVID-19 infection or exposure to COVID-19. These follow the typical states of the SEIR model, with additional levels of infectiousness included, as can be observed in Supplementary Materials C.5. Agents are able to move from one health state to another through the pathways indicated and are not required to go through every health state before they recover.

Covasim has many parameters predefined with a default value, including the probabilities associated with onward transmission and disease progression, duration of disease burden and the effects of interventions. These have been regularly updated as and when new evidence has become available since Covasim's development in 2020. Covasim is pre-populated with demographic data on population age structures and household sizes by country, and uses these data to generate its population contact network. In this work, we have used the demographic data and population network for the UK, although our modelling focuses on England only. Covasim generates four different contact networks: schools, workplaces, households, and community settings. Each agent has an age-dependent fixed number of interactions with other agents within each contact layer per time step. These fixed numbers are drawn from a Poisson distribution, which we set as having a mean of 3 for households, 20 for schools, 20 for work and 20 for the community.

Within the model there are probabilities associated with moving from one health state to another, as described in Supplementary Materials D.4. These probabilities are age dependent; we know that susceptibility to infection and the likely progression of COVID-19 symptoms within an individual varies drastically with age. The per-contact transmission probability ($\beta$) that an infectious individual transmits the virus to a susceptible individual is assumed to depend on the contact network and intervention measures in place.

We model the three national lockdowns by simulating a reduction in frequency of contact across different layers of society, using Google mobility data https://ourworldindata.org/covid-mobility-trends, and by reducing the value of the parameter $\beta$. This is analogous to previous work.

Transmission in schools is simulated as being reduced by 86% during the third national lockdown (04/01/2021 - 08/03/2021), by 37% following this to the end of the first term of the 2021-2022 school year, and by 80% for the rest of the 2021-2022 school year and then by 90% onwards. Precise changes to the household, school, workplace and community $\beta$ can be seen in Supplementary Materials D.5.

We model different test-trace-isolate (TTI) strategies analogous to the policies in England over 2020-2023. From March 23, 2020, the policy was to test people presenting with severe COVID-19 symptoms and ask them to self-isolate and, starting on June 1, 2020, this approach was complemented with contact-tracing of those people testing positive for infection. This was relaxed later on in the epidemic. Covasim accounts for testing strategies via parameters that determine the probabilities with which symptomatic and non-symptomatic people receive a test each day. The daily probabilities were chosen by comparing the % of people tested over different points of the epidemic and can be seen in Supplementary Materials D.6. We assumed 100%

sensitivity and specificity of the testing, a delay of 1 day to receive the test result, and that individuals testing positive would immediately be isolated for 14 days. In the model, this isolation reduced their infectiousness by varying levels, representing fluctuating levels of resistance to follow isolation guidance within the population (see Supplementary Materials D.7 for details).

Contract tracing that was part of the NHS Test and Trace scheme is also modelled. The effectiveness of this contact tracing method is derived via three variables: the probability that a contact can be traced, the time taken to trace the contact, and the decision that an individual makes once they have been traced. We model that an individual has a contact layer dependent probability of being traced, so that community layer contacts, for example, are less likely to be traced than household contacts. We also assume that some contacts take longer to trace than others, so there is a delay when tracing those in the community, workplace or school. A contact traced individual then has a probability associated with how likely they are to comply with isolation, as well as a duration for how long they are likely to isolate for. Further details of the methods used can be found in Supplementary Materials D.8.

Similarly to our previous work, we model an age-prioritised vaccination schedule with two vaccine doses given 8–12 weeks apart. Vaccines provide partial protection against SARS-CoV-2 infection indirectly. Vaccination reduces the likelihood that an individual: becomes infected once exposed; becomes symptomatic once infected; and becomes more severely ill once symptomatic. This represents what has been observed in the literature: the vaccination provides some protection against those who have mild/asymptomatic infections as well as those who are symptomatic [30]. Through this symptomatic reduction, the vaccine offers an indirect protection against transmission. The vaccination schedule we use reflects the vaccination roll out in place in England over the period December 2020 - February 2023 https://coronavirus.data.gov.uk, and includes the use of the Pfizer/BioNTech vaccine for individuals aged 65+ or under 40, and the Oxford/AstraZeneca vaccine for individuals aged 40–64. We incorporated the vaccination of those under 18 years from late August 2021 in our Baseline scenario and our adolescent booster vaccine scenario. The details of the vaccination roll out can be found in Supplementary Materials D.9.

We include two booster vaccines which follow an age prioritised structure. Rather than using probabilities to determine the booster vaccine roll out, we input values for the daily number of boosters given, $n_{boost}(t)$, on day $t$. These values are explained in Supplementary Materials D.10. We simulate the roll out of a booster vaccine for all adults in 2021, and an additional booster vaccine roll out in spring 2022 to those aged over 75 and in the autumn 2022 for those aged over 50.

A key feature of Covasim, and where it stands out from other COVID-19 ABMs, is its ability to mechanistically model individual SARS-CoV-2 variants by allowing different model parameters to be introduced which characterise each variant (see Supplementary Materials D.11 and Supplementary Materials D.12). Extending previous work, we have modelled all the consecutive variants that were circulating in England between January 2020 and April 2023: the wild type of SARS-CoV-2 which emerged in early 2020, the B.177 variant which emerged in August 2020, the Alpha variant which emerged in late September 2020 and spread nationally between October 2020 and February 2021, the Delta variant which emerged in late April 2021 and spread nationally until September 2021 and the various Omicron variants that spread in England from late 2021 onwards.

Within Covasim the parameters that characterise these variants are (a) the number of seed infections for each emerging variant at the time of emergence, (b) the relative transmissibility and severity of each variant, (c) VE against each variant and (d) the cross-immunity between variants.

In this analysis, the number of imported cases, transmissibility and severity for each variant is determined during the calibration process. For the effectiveness of vaccines against earlier variants we used the default Covasim values with details in Supplementary Materials D.13. The cross-immunities between the wild type, Alpha, B.1.177 and Delta variants were determined during the calibration process. We continue this for the Omicron sub-variants too, but assume a higher level of cross-immunity between different Omicron variants due to there being a higher level of shared genetics between them. The overall values are derived in the calibration process so that the pattern of emergence and proportion of variants matches that seen in the genomic data from England from https://covid19.sanger.ac.uk/lineages/raw. Details of the cross-immunities and VE against each variant used here can be found in Supplementary Materials D.14 and Supplementary

Materials D.15 respectively.

We also need to adjust the mean time taken for an agent to move between different infectious states. Omicron variants were found to have a shorter incubation period so we amended the mean time taken for an agent to move from an exposed to infectious state on 07/11/2021, reflecting the majority of new infections from this time onwards being of Omicron lineage. Details are in Supplementary Materials D.16.

Our previous Covasim work models the epidemic from 20/01/2020 to 20/06/2021, including all major variants in England up to Delta. Here we extend this work to include all of the Delta period and also include transmission of Omicron and its subvariants, simulating the COVID-19 epidemic up in England until April 2023.

We calibrated the model to data from the UK-COVID-19 dashboard using Optuna's hyper-parametrisation framework which is integrated with Covasim. We found optimal values for: (a) the number of seeded infections of the different Omicron variants of concern; and (b) the relative transmissibility and severity of each of the Omicron variants compared to previous circulating variants.

### C Additional Figures

#### C.1 Estimated Proportion of Infections Belong to Each Variant

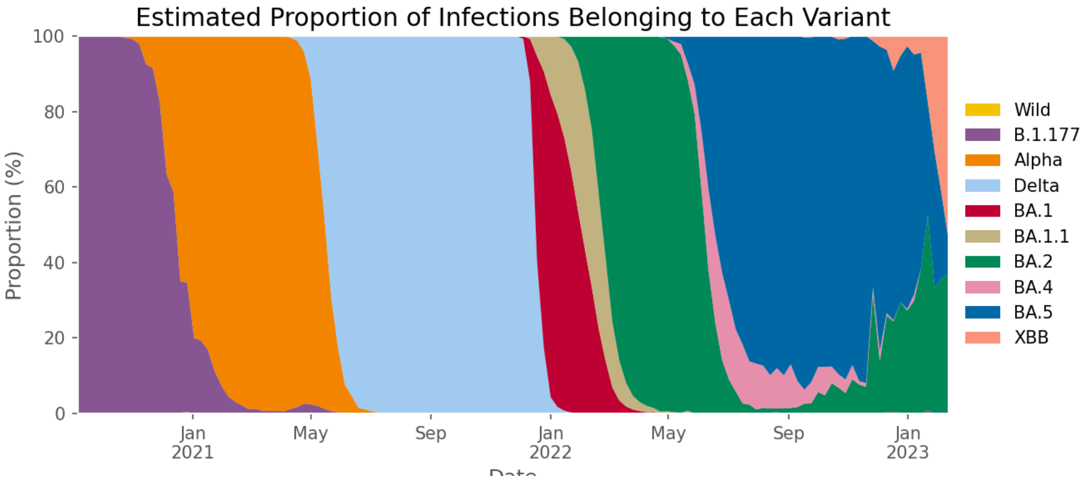


Figure 7: Estimated proportion of infections belonging to each variant between 05/09/2020 and 11/02/2023 in England. Data sourced from the Sanger Institute COVID-19 Genomic https://covid19.sanger.ac.uk/lineages/raw

#### C.2 Hospitalisations and Deaths for the Varying Adolescent Vaccine Uptake Scenarios

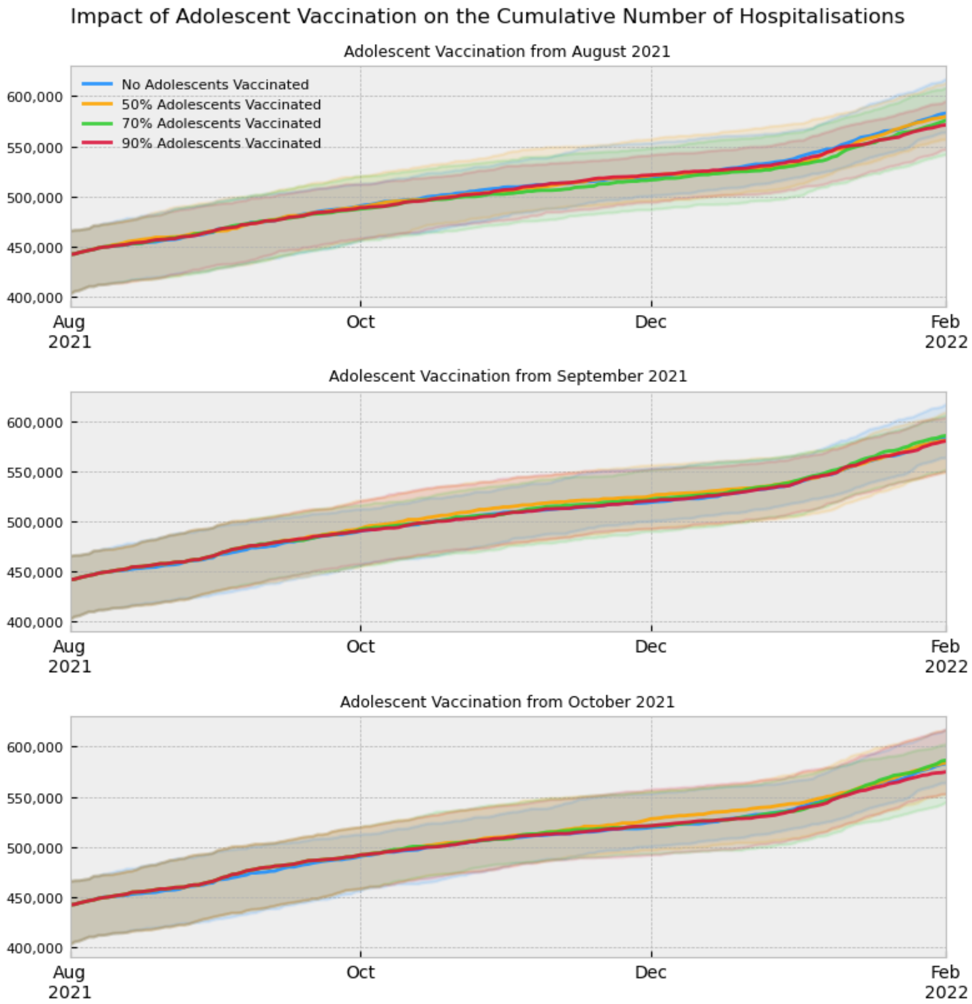


Figure 8: Comparative graphs showing the impact of varying adolescent vaccine uptake on the impact of the COVID-19 epidemic in England through the number of hospitalisations in the whole population and how this changes when vaccination is started in early August, September or October 2021.

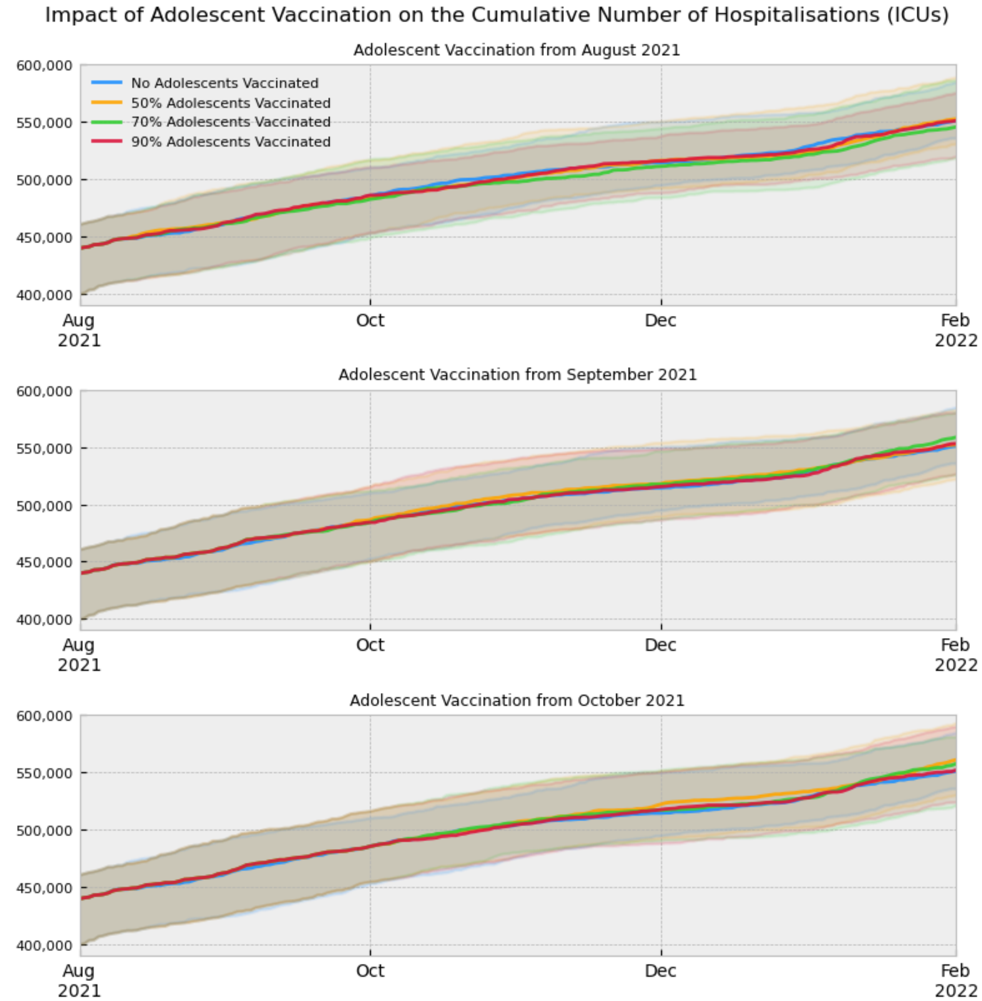


Figure 9: Comparative graphs showing the impact of varying adolescent vaccine uptake on the impact of the COVID-19 epidemic in England through the number of hospitalisations (ICUs) in the whole population and how this changes when vaccination is started in early August, September or October 2021.

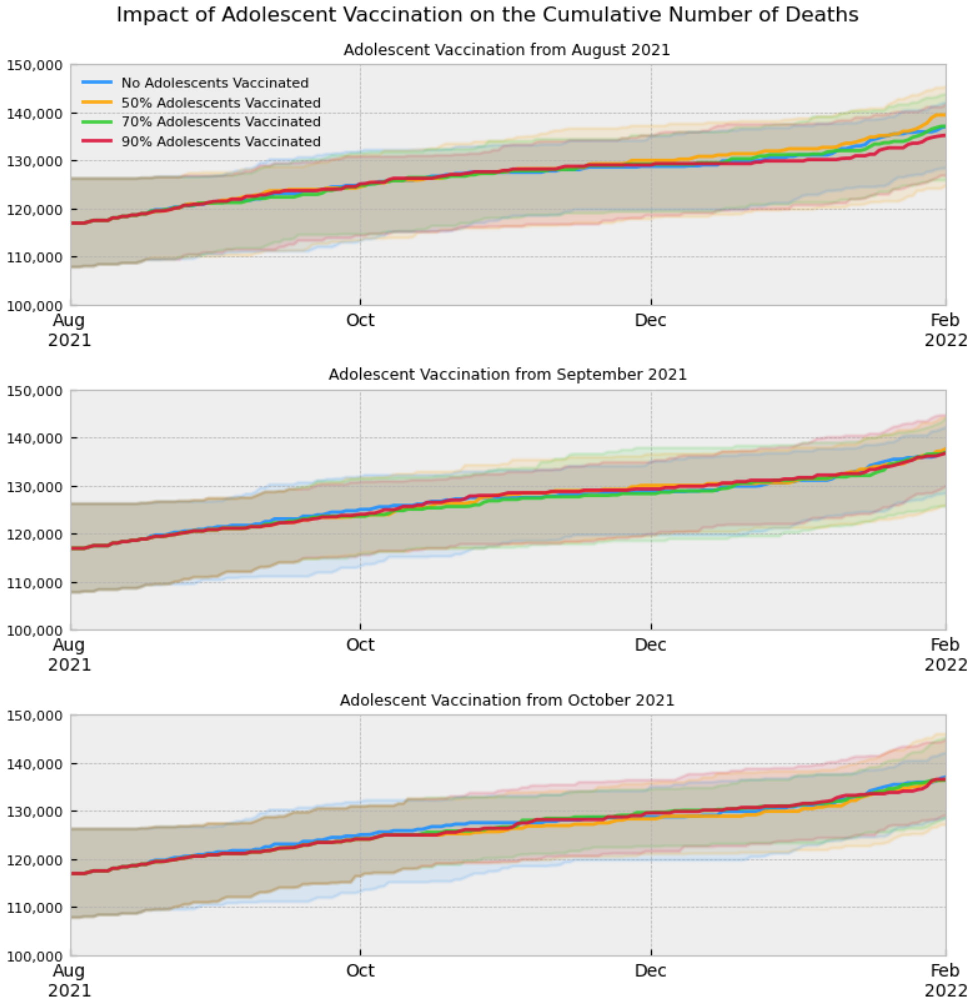


Figure 10: Comparative graphs showing the impact of varying adolescent vaccine uptake on the impact of the COVID-19 epidemic in England through the number of deaths in the whole population and how this changes when vaccination is started in early August, September or October 2021.

##### C.3 Adolescent Vaccination Uptake 3D Slice

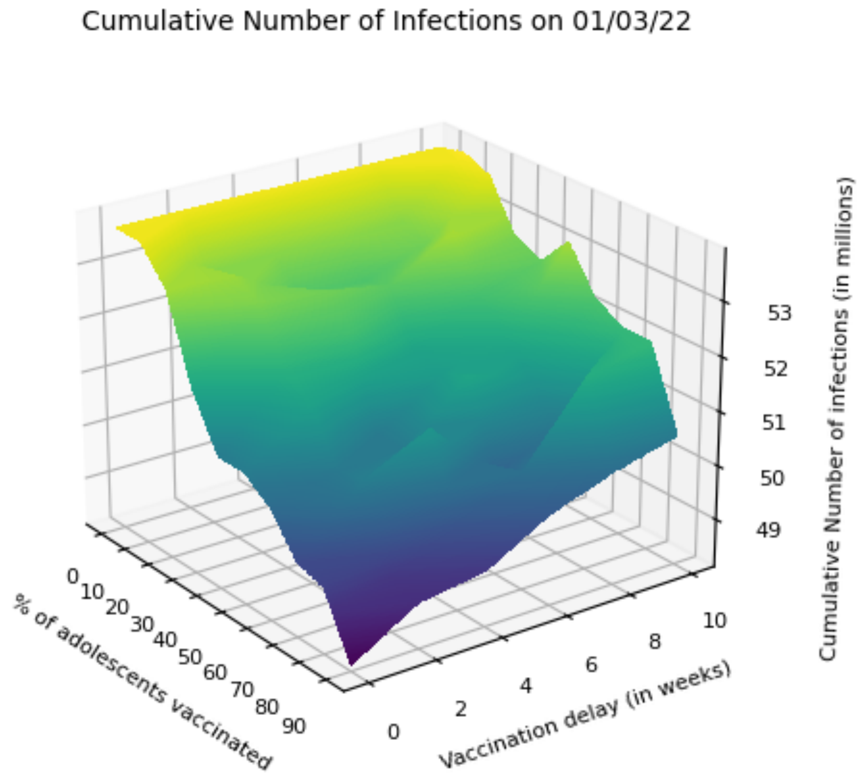


Figure 11: Cumulative number of infections on March, 01, 2022 under different scenarios of vaccine uptake and speed of vaccine roll out of immunisation against adolescents from August 2021.

#### C.4 Hospitalisations and Deaths for the Adolescent Booster Vaccine Scenario

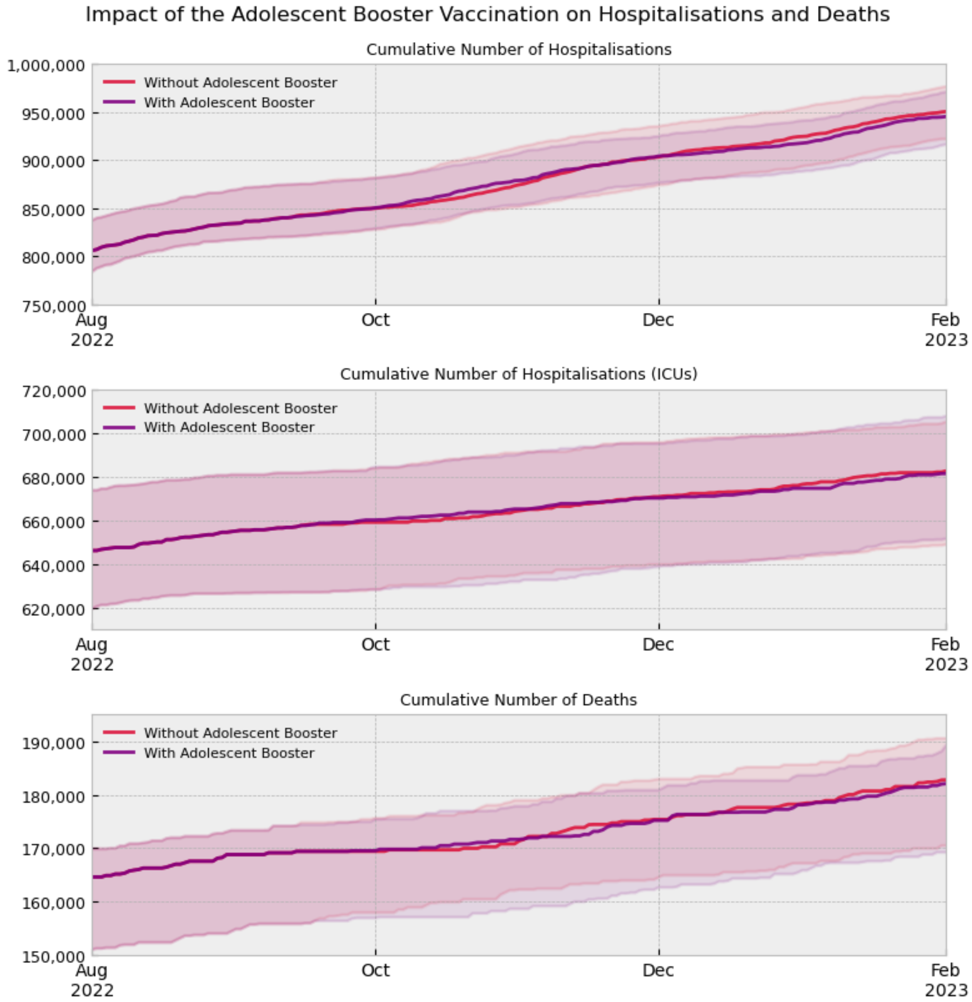


Figure 12: Comparative graphs showing the impact of a single dose of the booster vaccine for adolescents starting in September 2022 on the number of hospitalisations and deaths in the whole population.

#### C.5 Agent Health State Flow Chart

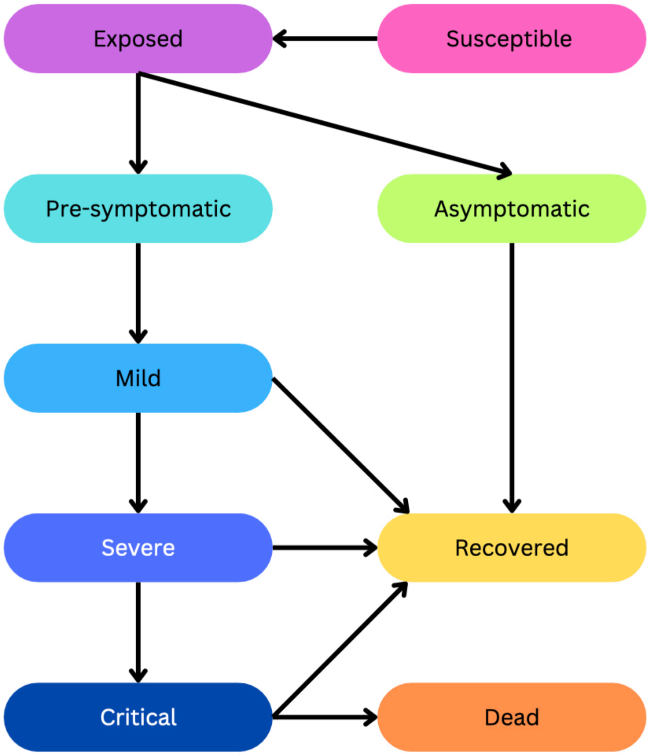


Figure 13: Flow chart indicating the main states that an agent can belong to and the possible routes that an agent's health can follow in Covasim. Each agent will belong to precisely one of these states at any one time. In addition, an agent may also be contact traced or tested which may then lead to them being quarantined or isolated. An agent is quarantined or isolated in addition to being in one of the main health states indicated in this flow chart.

### D Parameter Values

#### D.1 Daily Number of Infectious People

The data in the COVID-19 Infection Survey (https://www.ons.gov.uk) are given as a weekly average of the % of the population of England who were infectious. To convert this into the number of newly infectious people we use the following formula:

$$n_{daily}(t) = \frac{n_{weekly\%}(t) * 56\,000\,000}{7}, \tag{1}$$

where $n_{daily}(t)$ is the number of infectious people on day $t$ and $n_{weekly\%}(t)$ is the number of newly infectious people in the week spanned by day $t-3$ to $t+3$.

#### D.2 Adolescent Vaccination Uptake Scenarios

The following table shows how the approximate vaccine uptake in the adolescent vaccination scenarios is simulated. It shows the daily probability of vaccination and number of days the probability is simulated for, per age group (16-17 and 12-15). We assume that adolescents are given their second dose of the vaccine 12 weeks after their first dose. The lines in bold are the levels of adolescent vaccine uptake used for the main scenario analysis, and the rest are used only in the 3D slice plot.

We model the adolescent vaccination starting from different roll out dates. These are defined as a) August 2021: 01/08/2021 for ages 16-17 and 29/08/2021 for ages 12-15; b) September 2021: 29/08/2021 for ages 16-17 and 26/09/2021 for ages 12-15; and c) October 2021: 10/10/2021 for ages 16-17 and 07/11/2021 for ages 12-15.

The “days to reach" parameters were chosen to reflect realistic vaccination scenarios that are extensions of how fast vaccines were rolled out amongst adolescents. Higher uptake is assumed to have a longer rollout to reflect real-world limitations of availability of appointments for vaccines and the vaccines themselves. The “daily probability" parameter were calibrated to ensure that the approximate vaccine uptake was achieved over the days to reach allocated for each scenario.

| Approximate Final Uptake (%) | Age | Days to Reach | Daily Probability |
|---|---|---|---|
| **0** | **16-17** | **50** | **0** |
| | **12-15** | **71** | **0** |
| 10 | 16-17 | 60 | 0.0022 |
| | 12-15 | 81 | 0.0014 |
| 20 | 16-17 | 70 | 0.0032 |
| | 12-15 | 91 | 0.0025 |
| 30 | 16-17 | 80 | 0.0044 |
| | 12-15 | 101 | 0.0033 |
| 40 | 16-17 | 90 | 0.0058 |
| | 12-15 | 111 | 0.0046 |
| **50** | **16-17** | **100** | **0.0070** |
| | **12-15** | **121** | **0.0056** |
| 60 | 16-17 | 110 | 0.0081 |
| | 12-15 | 131 | 0.0070 |
| **70** | **16-17** | **120** | **0.0120** |
| | **12-15** | **141** | **0.0080** |
| 80 | 16-17 | 130 | 0.0125 |
| | 12-15 | 151 | 0.0105 |
| **90** | **16-17** | **140** | **0.0160** |
| | **12-15** | **161** | **0.0120** |

#### D.3 Adolescent Booster Vaccination Uptake Scenario

The adolescent booster vaccine scenario used the same vaccination as the baseline scenario, with the addition that 10,000 adolescents were vaccinated daily between 01/09/2022 and 31/12/2022. This was chosen to simulate a realistic number of adolescents being boosted (a similar proportion to the number of adolescents who were fully vaccinated in the baseline scenario) in a manageable time frame.

#### D.4 Predefined Disease Progression and Susceptibility Probabilities

The following table shows the age specific disease progression and susceptibility probabilities. $r_{sus}$ is the relative susceptibility an individual has to infection. $p_{sym}$, $p_{sev}$, $p_{cri}$, $p_{dea}$ are the probabilities that an individual develops mild, severe or critical symptoms or dies, respectively. See [19] for details of why these parameters have been predefined this way.

| | Age | | | | | | | | | |
|---|---|---|---|---|---|---|---|---|---|---|
| | 0-9 | 10-19 | 20-29 | 30-39 | 40-49 | 50-59 | 60-69 | 70-79 | 80-89 | 90+ |
| $r_{sus}$ | 0.34 | 0.67 | 1.00 | 1.00 | 1.00 | 1.00 | 1.00 | 1.24 | 1.47 | 1.47 |
| $p_{sym}$ | 0.50 | 0.55 | 0.60 | 0.65 | 0.70 | 0.75 | 0.80 | 0.85 | 0.90 | 0.90 |
| $p_{sev}$ | 0.00050 | 0.00165 | 0.00720 | 0.02080 | 0.03430 | 0.07650 | 0.13280 | 0.20655 | 0.24570 | 0.24570 |
| $p_{cri}$ | 0.00003 | 0.00008 | 0.00036 | 0.00104 | 0.00216 | 0.00933 | 0.03639 | 0.08923 | 0.17420 | 0.17420 |
| $p_{dea}$ | 0.00002 | 0.00002 | 0.00010 | 0.00032 | 0.00098 | 0.00265 | 0.00766 | 0.02439 | 0.08292 | 0.16190 |

#### D.5 Contact Layer Transmissibility

The following table gives the factors by which the $\beta$ values of each contact layer are multiplied by at different times to represent behavioural changes over the course of the pandemic. These parameters were found through calibration of the Baseline Scenario using Optuna, and reflected Google mobility data.

| Date | | Factor by which $\beta$ is multiplied | | | |
|---|---|---|---|---|---|
| Start | End | Household | School | Work Place | Community |
| 20/01/2020 | 14/02/2020 | 1.00 | 1.00 | 1.00 | 1.00 |
| 14/02/2020 | 16/03/2020 | 1.00 | 1.00 | 0.90 | 0.90 |
| 16/03/2020 | 23/03/2020 | 1.00 | 0.90 | 0.80 | 0.80 |
| 23/03/2020 | 01/06/2020 | 1.00 | 0.02 | 0.20 | 0.20 |
| 01/06/2020 | 15/06/2020 | 1.00 | 0.23 | 0.40 | 0.40 |
| 15/06/2020 | 22/07/2020 | 1.00 | 0.38 | 0.50 | 0.50 |
| 22/07/2020 | 29/07/2020 | 1.15 | 0.00 | 0.30 | 0.50 |
| 29/07/2020 | 02/09/2020 | 1.15 | 0.00 | 0.30 | 0.70 |
| 02/09/2020 | 26/10/2020 | 1.15 | 0.63 | 0.50 | 0.70 |
| 26/10/2020 | 05/11/2020 | 1.15 | 0.00 | 0.50 | 0.70 |
| 05/11/2020 | 14/11/2020 | 1.15 | 0.63 | 0.30 | 0.30 |
| 14/11/2020 | 21/11/2020 | 1.15 | 0.63 | 0.30 | 0.40 |
| 21/11/2020 | 10/12/2020 | 1.15 | 0.63 | 0.40 | 0.50 |
| 10/12/2020 | 24/12/2020 | 1.50 | 0.63 | 0.40 | 0.70 |
| 24/12/2020 | 04/01/2021 | 1.50 | 0.00 | 0.40 | 0.70 |
| 04/01/2021 | 15/02/2021 | 1.00 | 0.14 | 0.20 | 0.30 |
| 15/02/2021 | 22/02/2021 | 1.00 | 0.00 | 0.20 | 0.30 |
| 22/02/2021 | 08/03/2021 | 1.00 | 0.14 | 0.20 | 0.30 |
| 08/03/2021 | 29/03/2021 | 1.05 | 0.63 | 0.20 | 0.25 |
| 29/03/2021 | 12/04/2021 | 1.05 | 0.00 | 0.25 | 0.25 |
| 12/04/2021 | 19/04/2021 | 1.05 | 0.00 | 0.30 | 0.25 |
| | | | | | Continued on next page |

| Date | | Factor by which $\beta$ is multiplied | | | |
|---|---|---|---|---|---|
| Start | End | Household | School | Work Place | Community |
| 19/04/2021 | 17/05/2021 | 1.05 | 0.63 | 0.30 | 0.30 |
| 17/05/2021 | 31/05/2021 | 1.05 | 0.63 | 0.30 | 0.50 |
| 31/05/2021 | 07/06/2021 | 1.05 | 0.00 | 0.30 | 0.50 |
| 07/06/2021 | 19/06/2021 | 1.05 | 0.63 | 0.30 | 0.50 |
| 19/06/2021 | 28/06/2021 | 1.05 | 0.63 | 0.40 | 0.70 |
| 28/06/2021 | 19/07/2021 | 1.30 | 0.63 | 0.40 | 1.40 |
| 19/07/2021 | 07/09/2021 | 1.05 | 0.00 | 0.40 | 0.60 |
| 07/09/2021 | 22/10/2021 | 1.05 | 0.80 | 0.30 | 0.50 |
| 22/10/2021 | 05/11/2021 | 1.05 | 0.00 | 0.30 | 0.50 |
| 05/11/2021 | 01/12/2021 | 1.05 | 0.63 | 0.30 | 0.50 |
| 01/12/2021 | 09/12/2021 | 1.20 | 0.63 | 0.30 | 0.60 |
| 09/12/2021 | 16/12/2021 | 1.40 | 0.63 | 0.40 | 0.70 |
| 16/12/2021 | 20/12/2021 | 1.60 | 0.63 | 0.40 | 0.70 |
| 20/12/2021 | 04/01/2022 | 2.20 | 0.00 | 0.40 | 1.50 |
| 04/01/2022 | 30/01/2022 | 1.00 | 0.80 | 0.40 | 0.60 |
| 30/01/2022 | 15/02/2022 | 1.00 | 0.80 | 0.50 | 0.60 |
| 15/02/2022 | 22/02/2022 | 1.00 | 0.00 | 0.50 | 0.70 |
| 22/02/2022 | 09/04/2022 | 1.00 | 0.90 | 0.50 | 0.70 |
| 09/04/2022 | 22/04/2022 | 1.00 | 0.00 | 0.50 | 0.70 |
| 22/04/2022 | 28/05/2022 | 1.00 | 0.90 | 0.50 | 0.70 |
| 28/05/2022 | 03/06/2022 | 1.00 | 0.00 | 0.50 | 0.70 |
| 03/06/2022 | 23/07/2022 | 1.00 | 0.90 | 0.50 | 0.90 |
| 23/07/2022 | 31/08/2022 | 1.00 | 0.00 | 0.40 | 0.50 |
| 31/08/2022 | 22/10/2022 | 1.00 | 0.90 | 0.50 | 0.50 |
| 22/10/2022 | 29/10/2022 | 1.00 | 0.00 | 0.50 | 0.50 |
| 29/10/2022 | 19/12/2022 | 1.00 | 0.90 | 0.50 | 0.60 |
| 19/12/2022 | 04/01/2023 | 2.40 | 0.00 | 0.50 | 1.30 |
| 04/01/2023 | 30/04/2023 | 1.00 | 0.90 | 0.50 | 0.70 |

#### D.6 Testing Probabilities

The following table gives the daily probability that an asymptomatic/symptomatic agent is tested in our simulation. Once an agent has been tested, there is a 1 day delay to them receiving the test result. These parameters were found through approximate calibration of the Baseline Scenario to testing data for England. Calibration using Optuna was not appropriate as the nature of testing varied throughout the pandemic.

| Date | | Daily Testing Probability | |
|---|---|---|---|
| Start | End | Asymptomatic | Symptomatic |
| 20/01/2020 | 16/03/2020 | 0.00000 | 0.00000 |
| 16/03/2020 | 01/04/2020 | 0.00000 | 0.00900 |
| 01/04/2020 | 01/05/2020 | 0.00000 | 0.01200 |
| 01/05/2020 | 01/06/2020 | 0.00076 | 0.01200 |
| 01/06/2020 | 01/08/2020 | 0.00076 | 0.04769 |
| 01/08/2020 | 01/09/2020 | 0.00280 | 0.04769 |
| 01/09/2020 | 01/11/2020 | 0.00280 | 0.07769 |
| 01/11/2020 | 01/12/2020 | 0.00400 | 0.07769 |
| 01/12/2020 | 01/02/2021 | 0.00630 | 0.07769 |

Continued on next page

| Date | | Daily Testing Probability | |
|---|---|---|---|
| Start | End | Asymptomatic | Symptomatic |
| 01/02/2021 | 08/03/2021 | 0.00630 | 0.06769 |
| 08/03/2021 | 20/06/2021 | 0.00800 | 0.08769 |
| 20/06/2021 | 10/07/2021 | 0.02000 | 0.19769 |
| 10/07/2021 | 19/07/2021 | 0.01600 | 0.19769 |
| 19/07/2021 | 20/09/2021 | 0.00800 | 0.04769 |
| 20/09/2021 | 22/10/2021 | 0.00800 | 0.06769 |
| 22/10/2021 | 07/11/2021 | 0.00400 | 0.06769 |
| 07/11/2021 | 01/12/2021 | 0.00800 | 0.07769 |
| 01/12/2021 | 10/12/2021 | 0.01600 | 0.12000 |
| 10/12/2021 | 05/01/2022 | 0.02000 | 0.15000 |
| 05/01/2022 | 01/02/2022 | 0.00800 | 0.10000 |
| 01/02/2022 | 24/02/2022 | 0.00800 | 0.03000 |
| 24/02/2022 | 20/03/2022 | 0.00400 | 0.03000 |
| 20/03/2022 | 01/04/2022 | 0.00400 | 0.02000 |
| 01/04/2022 | 30/04/2023 | 0.00080 | 0.00800 |

#### D.7 Isolation Transmissibility

The following table gives the factors by which the $\beta$ values of an agent are multiplied by at different times to represent varying levels of compliance with isolation. These values override the default values Covasim uses for this which are given in Supplementary Materials D.4. These parameters were found through calibration of the Baseline Scenario using Optuna, and reflect changes in the requirements and recommendations for isolation at different points during the pandemic.

| Date | | Factor by which $\beta$ is multiplied by when in isolation |
|---|---|---|
| Start | End | |
| 01/04/2020 | 01/07/2020 | 0.2 |
| 01/07/2020 | 01/09/2020 | 0.4 |
| 01/09/2020 | 01/11/2020 | 0.6 |
| 01/11/2020 | 01/12/2020 | 0.2 |
| 01/12/2020 | 20/06/2021 | 0.5 |
| 20/06/2021 | 02/08/2021 | 0.8 |
| 02/08/2021 | 20/09/2021 | 0.2 |
| 20/09/2021 | 01/12/2021 | 0.6 |
| 01/12/2021 | 24/02/2022 | 0.7 |
| 24/02/2022 | 01/01/2023 | 0.8 |

#### D.8 Contract Tracing and Isolation Probability

The probability that an agent is traced needs to be varied to ensure that the diagnoses and infections match up to the observed data. However, the time it takes to trace an individual remains constant. The implemented contact tracing has the following structure: An individual is contact traced after $t_*$ days with probability $p_*$, where $*$ is one of $h, s, w$ or $c$ representing an interaction in a household, school, work or community layer, respectively. The values of $t_*$ are consistently

$$t_h = 0, \qquad t_s = 1, \qquad t_w = 1, \qquad t_c = 2.$$

The values of $p_*$ are

$$p_h = 1.0, \qquad p_s = 0.8, \qquad p_w = 0.8, \qquad p_c = 0.1.$$

We vary the duration, $\tau$, which contact traced individuals quarantine for. The values of $\tau$ are

$$\tau = \begin{cases} 10 & \text{if } 01/06/2020 \leq t < 10/09/2021, \\ 7 & \text{if } 10/09/2021 \leq t < 01/04/2022, \\ 5 & \text{if } 01/04/2022 \leq t < 30/04/2023. \end{cases} \tag{2}$$

These values were chosen to reflect realistic timelines and likelihoods of individuals being contact traced, dependent on which contact layer they were being contact traced through (household, school, work or community). The quarantine duration parameters reflect changes in the requirements and recommendations for quarantine at different points during the pandemic.

#### D.9 Vaccination Uptake

The following table gives the final approximate vaccine uptake in the Baseline Scenario as a proportion of each age group. It includes the first day of vaccination, the daily probability of being vaccinated and the number of days this probability is simulated for, per age group. The “start day" and “days to reach" reflect how the vaccine was rolled out in England. The “daily probability" is chosen so that the “approximate final uptake" matches that observed in England.

Those between the ages of 18 and 80 are assumed to be given their two doses of the vaccine at an interval of 8 weeks apart. All other vaccinated age groups are given their second dose 12 weeks after the first dose. This vaccination schedule is kept constant in all scenarios, with the exception of those aged 16-17 and 12-15 for the varying adolescent vaccination scenarios in the autumn of 2021 (see Supplementary Materials D.2 for further details).

| Age | Start Day | Approximate Final Uptake (%) | Days to Reach | Daily Probability |
|---|---|---|---|---|
| 90-99 | 08/12/2020 | 94.2 | 41 | 0.0750 |
| 85-89 | 08/12/2020 | 95.9 | 41 | 0.0750 |
| 80-84 | 08/12/2020 | 95.9 | 41 | 0.0750 |
| 75-79 | 18/01/2021 | 95.6 | 19 | 0.1500 |
| 70-74 | 29/01/2021 | 93.9 | 18 | 0.1500 |
| 65-69 | 15/02/2021 | 91.9 | 15 | 0.1620 |
| 60-64 | 01/03/2021 | 90.3 | 16 | 0.1360 |
| 55-59 | 06/03/2021 | 88.5 | 15 | 0.1350 |
| 50-54 | 17/03/2021 | 85.8 | 27 | 0.0710 |
| 45-49 | 13/04/2021 | 80.6 | 17 | 0.0930 |
| 40-44 | 26/04/2021 | 75.7 | 19 | 0.0740 |
| 35-39 | 13/05/2021 | 71.1 | 15 | 0.0810 |
| 30-34 | 20/05/2021 | 68.0 | 19 | 0.0578 |
| 25-29 | 08/06/2021 | 66.5 | 15 | 0.0705 |
| 18-24 | 15/06/2021 | 70.1 | 50 | 0.0240 |
| 16-17 | 23/08/2021 | 62.2 | 100 | 0.0086 |
| 12-15 | 20/09/2021 | 46.4 | 121 | 0.0051 |
| 5-11 | 01/03/2022 | 10.4 | 31 | 0.0034 |

#### D.10 Daily Booster Vaccines

Booster vaccines are rolled out in an age-prioritised way to those that are fully vaccinated. The cumulative number of booster vaccines simulated in the baseline strategy are given in Tables 9 to 11.

| Date, $\tau$ | Cumulative number of booster vaccines given, $C$ |
|---|---|
| 22/07/2021 | 0* |
| 22/08/2021 | 13 174 |
| 22/09/2021 | 117 995 |
| 22/10/2021 | 4 814 524 |
| 22/11/2021 | 12 799 122 |
| 22/12/2021 | 26 313 812 |
| 22/01/2022 | 30 375 660 |
| 22/02/2022 | 31 224 495 |
| 22/03/2022 | 31 659 900 |
| 22/04/2022 | 32 032 920 |
| 22/05/2022 | 32 361 679 |
| 22/06/2022 | 32 574 562 |
| 22/07/2022 | 32 758 281 |
| 22/08/2022 | 32 886 824 |
| 22/09/2022 | 32 972 761 |
| 22/10/2022 | 33 112 687 |
| 22/11/2022 | 33 226 445 |
| 01/01/2023 | 33 278 622 |

Table 9: This table shows the cumulative number of first dose booster vaccines given as part of the 2021 booster vaccine programme. *The correct value is 10 272, however, for calculation purposes, we use 0 so that we do not miss a section of booster vaccines in our model.

| Date, $\tau$ | Cumulative number of booster vaccines given, $C$ |
|---|---|
| 22/03/2022 | 0** |
| 22/04/2022 | 2 020 011 |
| 22/05/2022 | 3 394 681 |
| 22/06/2022 | 3 847 726 |
| 22/07/2022 | 4 029 669 |
| 22/08/2022 | 4 076 445 |
| 22/09/2022 | 4 080 680 |

Table 10: This table shows the cumulative number of second dose booster vaccines given to those aged over 75 as part of the spring 2022 booster vaccine programme. **The correct value is 42 676, however, for calculation purposes, we use 0 so that we do not miss a section of booster vaccines in our model.

| Date, $\tau$ | Cumulative number of booster vaccines given, $C$ |
|---|---|
| 01/09/2022 | 0 |
| 01/10/2022 | 4 406 560 |
| 01/11/2022 | 11 534 718 |
| 01/12/2022 | 14 386 134 |
| 01/01/2023 | 14 885 148 |

Table 11: This table shows the cumulative number of second dose booster vaccines given to those aged over 50 as part of the autumn 2022 booster vaccine programme.

#### D.11 Variant Parameters

The following table shows the calibrated factors that the $\beta$, $p_{sym}$, $p_{sev}$, $p_{cri}$ and $p_{dea}$ of the original wild variant are multiplied by to get the transmissibility and likelihood of a change in an agent's infection stage for each new variant $y$. The values shown are used in all scenarios to find the relative transmissibility ($\beta$) and the relative probabilities of an agent moving from mild to severe infection ($p_{sev}$), severe to critical infection

($p_{cri}$) and critical infection to death ($p_{dea}$) for each new variant. These parameters were found through careful calibration of the Baseline Model using Optuna.

| | B.1.177 | Alpha | Delta | BA.1 | BA.1.1 | BA.2 | BA.4 | BA.5 | XBB |
|---|---|---|---|---|---|---|---|---|---|
| $y_\beta$ | 1.20 | 1.80 | 2.60 | 3.60 | 3.10 | 5.10 | 5.00 | 5.50 | 5.20 |
| $y_{p_{sev}}$ | 0.20 | 1.01 | 0.28 | 0.30 | 0.30 | 0.20 | 0.35 | 0.35 | 0.35 |
| $y_{p_{cri}}$ | 23.00 | 100.00 | 25.00 | 0.15 | 0.15 | 0.07 | 0.05 | 0.05 | 0.05 |
| $y_{p_{dea}}$ | 1.00 | 0.70 | 0.40 | 0.90 | 0.50 | 0.50 | 1.55 | 1.55 | 1.55 |

#### D.12 Predefined Variant Cross-Immunity

The following table presents the factors by which the immunity of an agent (against a particular variant) is adjusted by when they have been previously infected. The row corresponds to the variant they have previously been infected by, and the the column corresponds to the new variant they are exposed to. See [19] for details of why these parameters have been predefined this way.

| | | Variant | | | | |
|---|---|---|---|---|---|---|
| | | Wild | Alpha | Beta | Gamma | Delta |
| Variant | Wild | 1.000 | 0.500 | 0.500 | 0.340 | 0.374 |
| | Alpha | 0.500 | 1.000 | 0.800 | 0.800 | 0.689 |
| | Beta | 0.066 | 0.500 | 1.000 | 0.500 | 0.086 |
| | Gamma | 0.050 | 0.050 | 0.040 | 1.000 | 0.040 |
| | Delta | 0.374 | 0.689 | 0.086 | 0.088 | 1.000 |

#### D.13 Predefined Vaccine Immunity

The following table shows the factors by which the immunity of an agent against a particular variant is adjusted by when they have been vaccinated. The row corresponds to the variant they have previously been infected by, and the the column corresponds to the new variant they are exposed to. See [19] for details of why these parameters have been predefined this way.

| | | Variant | | | | |
|---|---|---|---|---|---|---|
| | | Wild | Alpha | Beta | Gamma | Delta |
| Vaccine | Pfizer | 1 | $\frac{1}{2}$ | $\frac{1}{10.3}$ | $\frac{1}{6.7}$ | $\frac{1}{2.9}$ |
| | Moderna | 1 | $\frac{1}{1.8}$ | $\frac{1}{4.5}$ | $\frac{1}{8.6}$ | $\frac{1}{2.9}$ |
| | AstraZeneca | 1 | $\frac{1}{2.3}$ | $\frac{1}{9}$ | $\frac{1}{2.9}$ | $\frac{1}{6.2}$ |
| | Johnson & Johnson | 1 | 1 | $\frac{1}{3.6}$ | $\frac{1}{3.4}$ | $\frac{1}{1.6}$ |
| | Novavax | 1 | $\frac{1}{1.12}$ | $\frac{1}{4.7}$ | $\frac{1}{8.6}$ | $\frac{1}{6.2}$ |
| | Sinovac | 1 | $\frac{1}{1.12}$ | $\frac{1}{4.7}$ | $\frac{1}{8.6}$ | $\frac{1}{6.2}$ |
| | Sinopharm | 1 | $\frac{1}{1.12}$ | $\frac{1}{4.7}$ | $\frac{1}{8.6}$ | $\frac{1}{6.2}$ |

#### D.14 Variant Cross-Immunity

The following table gives the factors by which the immunity of an agent is adjusted by when they have been previously infected by a specific variant of Covid-19. The row corresponds to the variant they have previously been infected by, and the column corresponds to the new variant they are exposed to. We only include the vaccines and variants used within our simulation. These parameters were found through calibration of the Baseline Scenario using Optuna.

| | | Variant | | | | | | | | | |
|---|---|---|---|---|---|---|---|---|---|---|---|
| | | Wild | B.1.177 | Alpha | Delta | BA.1 | BA.1.1 | BA.2 | BA.4 | BA.5 | XBB |
| Variant | Wild | 1.000 | 0.066 | 0.500 | 0.374 | 0.050 | 0.050 | 0.050 | 0.050 | 0.050 | 0.050 |
| | B.1.177 | 0.066 | 1.000 | 0.500 | 0.086 | 0.050 | 0.050 | 0.050 | 0.050 | 0.050 | 0.050 |
| | Alpha | 0.500 | 0.500 | 1.000 | 0.689 | 0.050 | 0.050 | 0.050 | 0.050 | 0.050 | 0.050 |
| | Delta | 0.374 | 0.086 | 0.689 | 1.000 | 0.040 | 0.040 | 0.040 | 0.040 | 0.040 | 0.040 |
| | BA.1 | 0.050 | 0.050 | 0.050 | 0.040 | 1.000 | 0.200 | 0.600 | 0.300 | 0.300 | 0.300 |
| | BA.1.1 | 0.050 | 0.050 | 0.050 | 0.040 | 0.200 | 1.000 | 0.600 | 0.200 | 0.200 | 0.100 |
| | BA.2 | 0.050 | 0.050 | 0.050 | 0.040 | 0.600 | 0.600 | 1.000 | 0.800 | 0.600 | 0.600 |
| | BA.4 | 0.050 | 0.050 | 0.050 | 0.040 | 0.100 | 0.200 | 0.800 | 1.000 | 0.500 | 0.400 |
| | BA.5 | 0.050 | 0.050 | 0.050 | 0.040 | 0.300 | 0.200 | 0.600 | 0.500 | 1.000 | 0.400 |
| | XBB | 0.050 | 0.050 | 0.050 | 0.040 | 0.300 | 0.100 | 0.600 | 0.400 | 0.400 | 1.000 |

#### D.15 Vaccine Effectiveness

This following table gives the factor by which the immunity of an agent is adjusted by when they are vaccinated. The row corresponds to the vaccine they received and the column corresponds to the new variant they are exposed to. We only include the vaccines and variants used within our simulation. These parameters were found through calibration of the Baseline Scenario using Optuna.

| | | Variant | | | | | | | | | |
|---|---|---|---|---|---|---|---|---|---|---|---|
| | | Wild | B.1.177 | Alpha | Delta | BA.1 | BA.1.1 | BA.2 | BA.4 | BA.5 | XBB |
| Vaccine | Pfizer | 1 | $\frac{1}{10.3}$ | $\frac{1}{2}$ | $\frac{1}{2.9}$ | $\frac{1}{4.5}$ | $\frac{1}{4.5}$ | $\frac{1}{4.5}$ | $\frac{1}{4.5}$ | $\frac{1}{4.5}$ | $\frac{1}{4.5}$ |
| | AstraZeneca | 1 | $\frac{1}{9}$ | $\frac{1}{2.3}$ | $\frac{1}{6.2}$ | $\frac{1}{2.9}$ | $\frac{1}{2.9}$ | $\frac{1}{2.9}$ | $\frac{1}{2.9}$ | $\frac{1}{2.9}$ | $\frac{1}{2.9}$ |
| | Booster* | 1 | $\frac{1}{9.6}$ | $\frac{1}{2.1}$ | $\frac{1}{4.5}$ | $\frac{1}{4.8}$ | $\frac{1}{4.8}$ | $\frac{1}{4.8}$ | $\frac{1}{4.8}$ | $\frac{1}{4.8}$ | $\frac{1}{4.8}$ |

*The Booster vaccine is a mix between the effectiveness of both the AstraZeneca and Pfizer vaccines as they were both used in England as booster vaccines. It is given as a single dose and boosts the level of NAbs by a factor of 3.

#### D.16 Incubation Period

The times taken to move between exposed to infectious (pre-symptomatic or asymptomatic) are known as viral shedding. We define $\tau_{xy}$ as the time taken to move from state $y$ to state $x$, where $x$ and $y$ are an allowed combination of ($a$, $e$, $p$, $s$, $m$, $c$, $d$, $r$), which represent asymptomatic, exposed, pre-symptomatic, severe, mild, critical, dead and recovered states respectively. The allowed combinations and default values are found in Supplementary Materials D.17. Once an agent has been exposed, there is a natural progression through the stages of infection which occurs over a number of time steps. Each subsequent viral shedding step is then modelled using a log-normal distribution with a specific mean $\mu$ and standard deviation $\sigma^2$.

We change the mean of $\tau_{ae}$, $\tau_{pe}$ and $\tau_{rs}$ to 3.4, 3.4 and 5.5, respectively, on 07/11/2021, as the majority of new infections from this time onwards were of Omicron lineage. The incubation period for different Covid-19 variants was fairly consistent up until the introduction of Omicron variants (autumn 2021). Omicron variants were found to have a shorter incubation period [31]. Hence, the required length of isolation time for infected individuals is shortened.

#### D.17 Predefined Distribution Parameters for $\tau_{xy}$

The following table shows the default mean and standard deviation associated with each log-normally distributed duration parameter $\tau_{xy}$. $\tau_{xy}$ represents the time taken to move from state $y$ to state $x$, where $x$

and $y$ are one of the allowed combinations of ($a$, $e$, $p$, $s$, $m$, $c$, $d$, $r$), which represent asymptomatic, exposed, pre-symptomatic, severe, mild, critical, dead and recovered states respectively. The allowed combinations are the ones seen in this table. See [19] for details of why these parameters have been predefined this way.

| Parameter | Health State Change | Mean | Standard Deviation |
|---|---|---|---|
| $\tau_{ae}$ | Exposed → Asymptomatic | 4.5 | 1.5 |
| $\tau_{pe}$ | Exposed → Pre-symptomatic | 4.5 | 1.5 |
| $\tau_{mp}$ | Pre-symptomatic → Mild | 1.1 | 0.9 |
| $\tau_{sm}$ | Mild → Severe | 6.6 | 4.9 |
| $\tau_{cs}$ | Severe → Critical | 1.5 | 2.0 |
| $\tau_{dc}$ | Critical → Dead | 10.7 | 4.8 |
| $\tau_{ra}$ | Asymptomatic → Recovered | 8.0 | 2.0 |
| $\tau_{rm}$ | Mild → Recovered | 8.0 | 2.0 |
| $\tau_{rs}$ | Severe → Recovered | 8.1 | 6.3 |
| $\tau_{rc}$ | Critical → Recovered | 8.1 | 6.3 |

#### D.18 Predefined Isolation and Quarantine Factors

The following table shows the factors by which each of an agent's contact layer $\beta$ values are multiplied by when they are in isolation or quarantine. For example, an (infectious) agent in isolation will have a 70% reduction to the probability that they transmit the virus when they come into contact with a susceptible agent in their household. See [19] for details of why these parameters have been predefined this way.

| | Factor by which $\beta$ is multiplied | | | |
|---|---|---|---|---|
| | Household | School | Work Place | Community |
| Isolation | 0.30 | 0.10 | 0.10 | 0.10 |
| Quarantine | 0.60 | 0.20 | 0.20 | 0.20 |
